# Decoding the genetic architecture of hernia through genome-wide association and multi-trait analyses

**DOI:** 10.64898/2026.05.12.26353033

**Authors:** Andrew M. Pregnall, Shuai Yuan, Jeremy M. Lawrence, Sarah A. Abramowitz, John DePaolo, Renae Judy, Gabrielle Shakt, Michael G. Levin, Scott M. Damrauer, Heather Wachtel

## Abstract

Hernias affect millions of individuals worldwide and represent a significant public health burden, yet the genetic mechanisms underlying hernia development and the extent to which they are shared across anatomical subtypes remains incompletely understood. We performed a multi-population genome-wide association meta-analysis of five hernia subtypes and identified 243 genome-wide significant loci, including 173 novel associations. Gene prioritization implicated genes involved in extracellular matrix organization, elastic fiber assembly, and embryologic development as key effectors of hernia susceptibility. Further analyses demonstrated substantial overlap in the genomic architecture of hernia, including 30 causal variants that were shared across different hernia subtypes. We employed genomic structural equation modeling to formally model this relationship, which identified two distinct latent genetic factors corresponding to putative midline fusion defects (ventral, umbilical, diaphragmatic) and inguinofemoral hernias (inguinal, femoral). Mendelian randomization analyses confirmed causal roles for body mass index, visceral adipose tissue, and abdominal subcutaneous adipose tissue in hernia development while also identifying candidate therapeutic targets. Together, these findings delineate the shared and distinct genetic architecture of hernia subtypes providing a mechanistic foundation to enable precision risk stratification and inform the development of novel preventative and therapeutic strategies.

## Introduction

Hernias are outpouchings of organs through defects in connective tissue. They occur in approximately 13 million patients annually and are increasing in prevalence.^1^ In 2019, more than 32 million patients were living with hernia; prevalence is projected to increase to 38 million patients by 2030, representing a significant public health burden.^2^ Clinically, hernias are subdivided by their anatomic location. Ventral, umbilical, and inguinal hernias are types of abdominal hernia. Ventral hernias occur through defects in the anterior abdominal wall; umbilical hernias occur through defects in the umbilicus; and inguinal hernias occur through defects in the inguinal canal. Femoral and diaphragmatic hernias are considered non-abdominal hernias, with femoral hernias occurring inferior to the inguinal ligament and diaphragmatic hernias occurring through defects in the diaphragm.

Observational studies have identified shared, unique and discordant risk factors in hernia subtypes, suggesting that, while some hernia mechanisms are conserved, others may be subtype-specific. Clinical risk factors for abdominal wall hernias include older age, smoking, and male sex.^3;4^ Although an increase in body mass index (BMI) is strongly associated with ventral, umbilical, and incisional hernias, a higher BMI paradoxically appears to be inversely associated with the risk of inguinal hernia.^5;6^

Similarly, population-based genetic studies have demonstrated that genetic susceptibility contributes to hernia development, including genetic collagenopathies such as Ehler-Danlos syndrome and Marfan syndrome^7–12^. These studies have identified a host of common susceptibility loci, but the extent to which genetic susceptibility to hernia is shared between subtypes remains incompletely understood. Several genome-wide association studies (GWAS) have further characterized genetic susceptibility to hernia. A recent UK Biobank study of five hernia subtypes (inguinal, diaphragmatic, umbilical, femoral and ventral) identified 26 loci shared across two or more hernia types, suggesting a common genetic mechanism.^13^ Additional UK Biobank analyses have identified loci linked to connective tissue, fibroblasts, and smooth muscle, offering further insight into hernia pathogenesis.^14;15^ However, prior studies have largely been limited to single biobanks, specific hernia subtypes, and European-only populations.

In this investigation, we conducted a multi-population GWAS meta-analysis of five hernia subtypes across five biobanks to identify novel susceptibility loci and to define the shared and distinct genetic architecture of hernia. Through Linkage-Disequilibrium Score Regression (LDSC), Local Analysis of Variance Association (LAVA), Bayesian colocalization analysis, and genomic structural equation modeling (Genomic-SEM), we define the shared and unique genetic architecture of hernia. Using Mendelian randomization, we elucidate causal phenotypes in hernia development, decode the causal role of increased BMI and adiposity traits in the development of inguinal hernia, and identify potential therapeutic targets for incisional hernia. Overall we report a comprehensive genomic analysis of hernia, establishing a biologically grounded framework for hernia risk stratification and identifying potential targets for hernia prevention and treatment.

## Results

### Multi-population meta-analysis identifies 243 loci across five hernia subtypes

The design of our study is presented in Figure 1. We performed multi-population GWAS meta-analyses for five hernia subtypes—ventral, inguinal, umbilical, femoral and diaphragmatic—using an inverse variance-weighted fixed-effects model on summary statistics from the All of Us Research Program,^16^ FinnGen,^17^ BioBank Japan,^18^ UK Biobank,^19^ and the VA’s Million Veteran Program.^20^ The meta-analysis included 23,354 people with ventral hernia and 1,596,564 without; 106,421 people with inguinal hernia and 1,729,615 without; 31,367 people with umbilical hernia and 1,581,741 without; 2,446 people with femoral hernia and 790,529 without; and 81,148 people with diaphragmatic hernia and 1,545,371 without (see Supplementary Data 1). Compared to the most recent GWAS of hernia, this represents a 5-fold increase in the effective sample size for ventral hernia, 2-fold increase in the effective sample size for inguinal hernia, 4.9-fold increase in the effective sample size for umbilical hernia, 2.3-fold increase in the effective sample size for femoral hernia, and 2.7-fold increase in the effective sample size for diaphragmatic hernia.^13^

**Figure 1:**
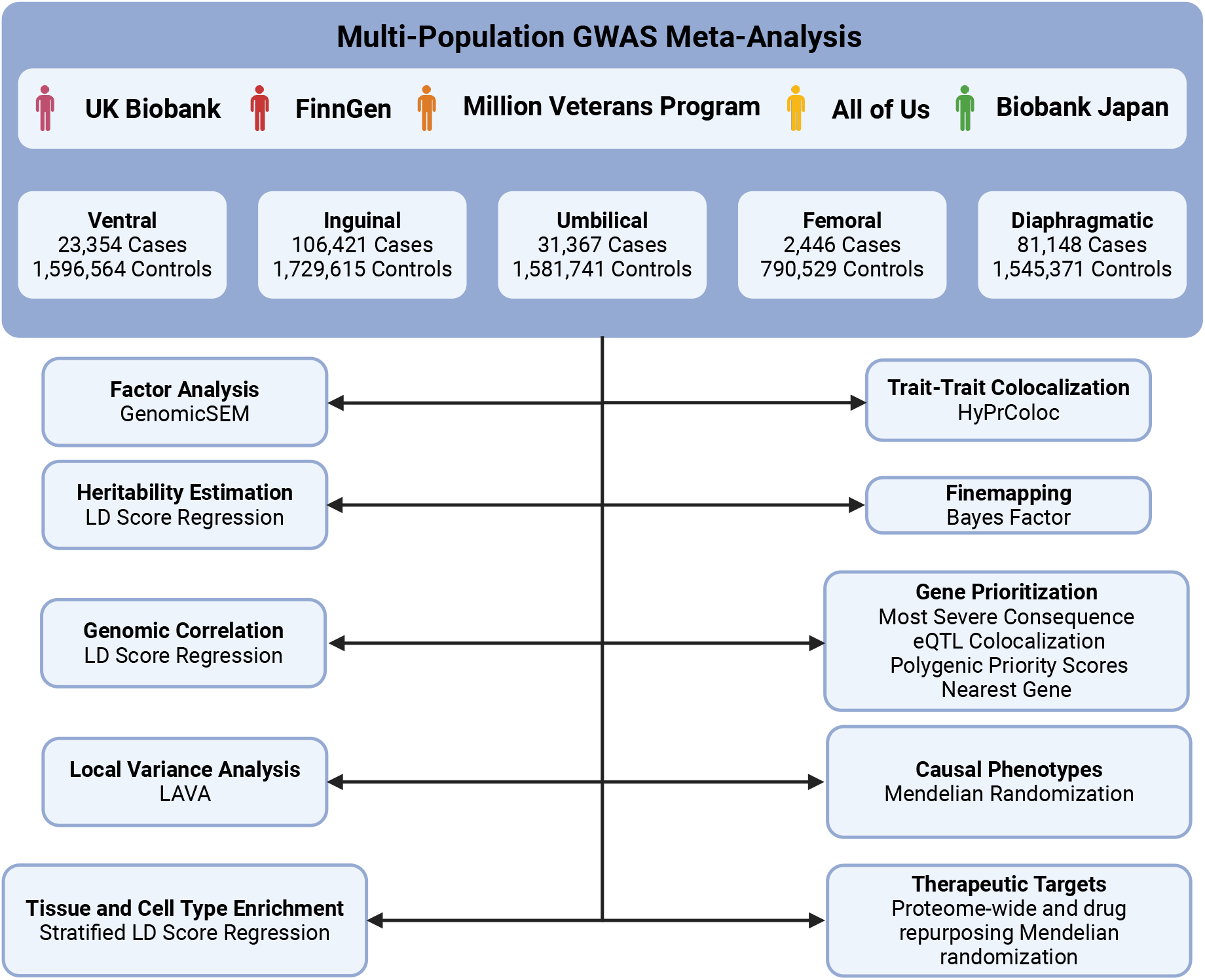
Overview of the study design and major analyses performed. Created in BioRender.

The analysis identified 30 loci associated with ventral hernia, 154 loci associated with inguinal hernia, 83 loci associated with umbilical hernia, 3 loci associated with femoral hernia, and 69 loci associated with diaphragmatic hernia which collapsed into 243 unique loci across all traits (see Figure 2, Supplementary Figure 1, and Supplementary Data 2-3). Of the 243 unique loci we identified, 70 overlapped with 1-Mb regions spanning previously reported lead variants (n = 80), providing replication for 87.5% of previously established loci. Given the multi-population nature of our cohort, we also performed meta-regression with MR-MEGA which partitions allelic effects across ancestral principal components; our meta-regression with MR-MEGA recapitulated all genome-wide significant loci results from the inverse variance weighted-based meta-regression, but did not identify any novel loci (see Supplementary Figure 3)

**Figure 2:**
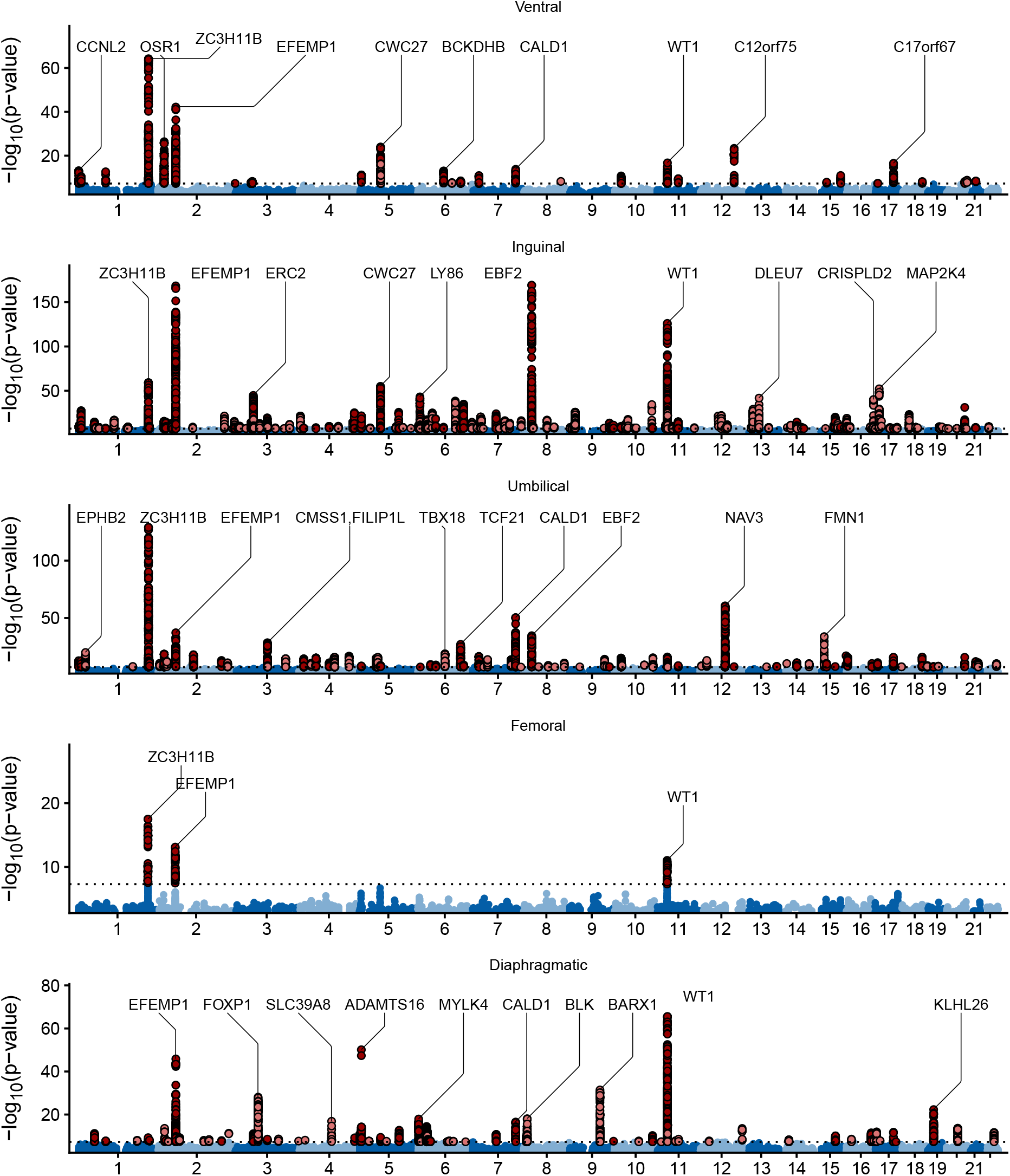
Manhattan plots from the inverse variance-weighted fixed effects meta-analysis of multi-population summary statistics for five hernia subtypes. Dark red dots indicate a genome-wide significant loci that is shared across at least two hernia subtypes. Light red dots indicate a genome-wide significant loci that is not shared across hernia subtypes. Significant loci are annotated with the nearest gene. Annotations are limited to the top ten most significant loci.

In addition to multi-population meta-analysis, we performed population-specific meta-analyses for each hernia subtype to identify population-specific genetic associations based on participant’s genetic similarity to the 1000 Genomes reference populations.^21^ Among individuals genetically similar to African reference populations, we identified one locus that reached genome-wide significance (*P <* 5 *×* 10^−8^) in the population-specific analysis but not in the multi-population or European-only meta-analyses. The lead variant for ventral hernia was 11:120719031:T:G (*β* = -0.2986, SE = 0.0545, *P* = 4.287 *×* 10^−8^). It has an minor allele frequency of 0.089 in the gnoMAD v4.1.1 African reference populations compared to an allele frequency of 0.049 in the European reference population (see Supplementary Figure 2). This variant is located nearest *GRIK4* which encodes a cation-permeable ligand-gated ion channel that functions as a glutamate receptor in central nervous tissues. Mouse models of *GRIK4* knockouts demonstrate increased total body fat and decreased lean body mass, suggesting increased or reduced mechanical tissue stress as a putative pathway through which this locus may modulate ventral hernia risk.^22^

### Integrative finemapping and functional annotation prioritize hernia causal variants and effector genes

We performed statistical finemapping using approximate Bayes factors to identify putatively causal genetic variants within genome-wide significant loci identified in our meta-analyses. In total, we finemapped 339 locus-trait pairs from the multi-population meta-analyses. The median size of the credible sets was 14 variants for ventral hernia, 17 variants for umbilical hernia, 33 variants for inguinal hernia, 40 variants for femoral hernia, and 39 variants for diaphragmatic hernia (see Figure 3A). In the multi-population meta-analysis, we identified 14 loci where only one variant was found in the 99% credible set, including several novel variant-trait associations at loci related to connective tissue structure and function. These variants included 15:62516340:C:T—an intronic variant of *TLN2* which is a component of focal adhesion plaques that mediates links between integrin and the actin cytoskeleton^23;24^—and 14:22848616:A:G located nearest MMP14 which is a peptidase that degrades extracellular matrix proteins including collagen and mediates actin cytoskeleton reorganization.^25;26^ Compared to genome-wide significant variants, single-variant credible sets were enriched for functional annotations (*P <* 0.001; Fisher’s exact test; see Figure 3B).

**Figure 3:**
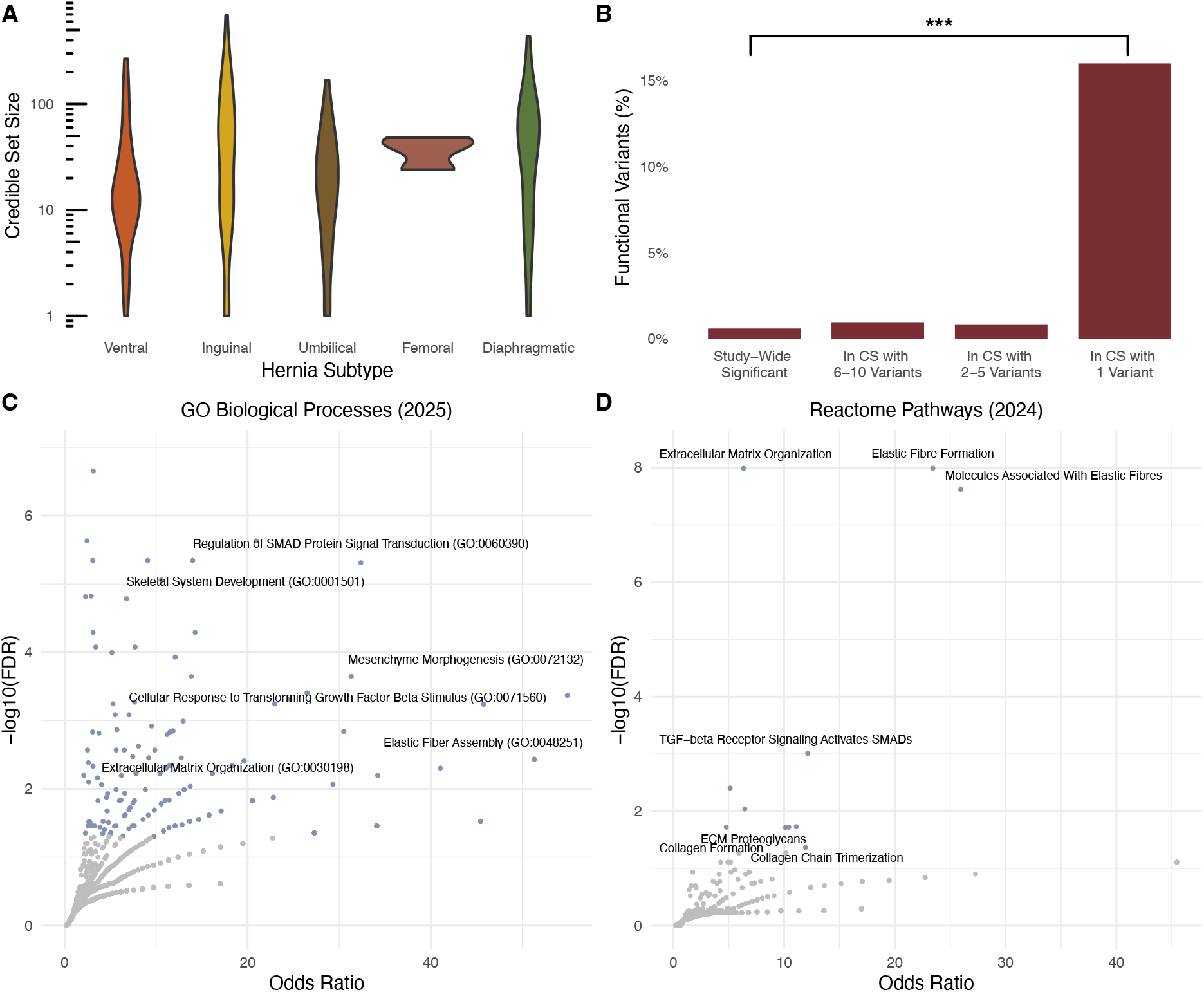
Multipopulation finemapping signals and variant-to-gene prioritization. (A) The distribution of credible set sizes, stratified by hernia subtype. (B) Proportion of functional variants, stratified by credible set size. (C) Gene set enrichment analysis of prioritized genes in the Go Biological Processes (2025). (D) Gene set enrichment analysis of prioritized genes in Reactome Pathways (2024).

To identify putatively causal genes associated with hernia risk, we used three complementary approaches: (1) variant consequence annotation via Ensembl Variant Effect Predictor (VEP),^27^ (2) expression quantitative trait loci (eQTL) colocalization and (3) Polygenic Priority Scores (PoPS).^28^ We prioritized genes meeting at least one of the following criteria: (1) the credible set contained a splice acceptor, splice donor, missense, or stop-gain variant as annotated by VEP; (2) there was a high posterior probability of colocalization between the trait and eQTL data for the cognate gene in fibroblasts (Posterior Probability for Colocalization (PPFC) > 0.7); or (3) the gene ranked within the top 10% of PoPS-prioritized genes among genome-wide significant loci for the phenotype. Genes at loci not meeting one of these orthogonal criteria were prioritized using the nearest gene approach. Using this approach, we prioritized 291 distinct genes of which 93 were prioritized using one or multiple orthogonal approaches (see Supplementary Data 5). Five novel hernia effector genes were prioritized through multiple lines of evidence. For diaphragmatic hernia, we prioritized *TRIOBP* and *UQCC1. TRIOBP* plays an essential role in actin cytoskeletal organization, cell spreading and contraction, and mitotic progression^29–32^ while UQCC1 is highly expressed in skeletal muscle tissue and fibroblasts and is required for the assembly of the ubiquinol-cytochrome c reductase complex—a key protein in aerobic metabolism.^33^ For inguinal hernia, we prioritized *TMEM204* which is highly expressed in adipose tissue and mediates cellular adhesion and permeability at adherans junctions,^34^ and *ZNF135* which is a regulator of cytoskeletal morphology and organization.^35^ Lastly, for umbilical hernia we prioritized *C1QTNF7* which is an adiponectin paralogue that is associated with metabolic derangements such as increased BMI and insulin resistance.^36–39^ In addition, *C1QTNF7* is predicted to be a component of collagen trimers, suggesting that this gene could contribute to hernia formation through both weakening of connective tissue and increased mechanical stress on connective tissue.

To formally assess biological pathways underlying shared hernia biology, we conducted a gene set enrichment analysis of prioritized genes using the GO Biological Process (2025) and Reactome Pathway (2024) databases (see Figure 3C/D and Supplementary Data 6-7). GO Biological Processes demonstrated enrichment for multiple pathways including “Regulation of SMAD Protein Signal Transduction”, “Cellular Response to TGF-*β* Stimulus”, “Mesenchyme Morphogenesis”, and “Elastic Fiber Assembly” highlighting the importance of growth signaling, embryonic processes, and extracellular matrix biology in hernia formation. Similarly, Reactome pathways demonstrated enrichment multiple pathways related to connective tissue biology including “Extracellular Matrix Organization”, “Molecules Associated With Elastic Fibres”, “Elastic Fibre Formation”, and “Collagen Formation”. Finally, eleven prioritized genes were shared across different hernia subtypes (*CCDC71, EBF2, FILIP1L, LRRK1, ERC2, QSOX2, SMURF2, ZNHIT6, EYA4, SPSB1*, and *OSR1*). These genes were enriched for biological processes related to embryological development including “Positive Regulation of Wnt Signaling Pathway”, “Embryonic Hindlimb Morphogenesis”, and “Positive Regulation of Gastrulation”, suggesting that common developmental biology pathways contribute to shared hernia susceptibility.

### Shared genomic architecture of hernia subtypes across global and local scales

Given the substantial overlap between lead loci identified in our GWAS meta-analysis and shared prioritized genes across traits, we sought to further elucidate the common genetic architecture of hernia subtypes. We applied cross-trait LDSC to estimate heritability of hernia subtypes. Heritability estimates ranged from 2.2-2.3% for diaphragmatic hernia to 17.9-24.9% for umbilical hernia (see Supplementary Figure 4). We also employed LDSC-SEG to partition hernia heritability across tissue and cell types. Enrichment patterns were pronounced in tissues consistent with connective tissue biology, including synovial and serosal membranes, as well as in adipose-related tissues comprising omentum and subcutaneous fat (see Supplementary Figure 6). At the cellular level, hernias demonstrated the strongest enrichment for signatures of mesenchymal-lineage and stromal cell types, including chondrocytes, mesenchymal stem cells, fibroblasts, and adipocytes (Supplementary Figure 7). Collectively, these enrichments provide biological context for the heritability of hernia, implicating mesenchymal/connective tissue programs and related stromal cell types in hernia susceptibility.

Using LDSC, we also estimated cross-trait genomic correlation between hernia subtypes. Genomic correlation coefficients ranged from 0.21–0.75, with the weakest association between diaphragmatic hernia and inguinal hernia (0.24) and the strongest association between ventral hernia and umbilical hernia (0.75) (see Figure 4A). Despite shared-liability and enrichment signals, genome-wide *r*_*g*_ estimates represent an average across the entire genome and can obscure locus-specific heterogeneity. To resolve whether shared architecture between subtypes is concentrated within specific genomic regions—and whether this sharing differs from the global *r*_*g*_ pattern—we next applied local genetic correlation analysis using LAVA.^40^ We restricted analyses to loci with sufficient SNP-based heritability for the disorders analyzed (*P <* 0.05*/*243); see Methods). We detected 267 significant pairwise local *r*_*g*_s (*P <* 0.05*/*961) after correcting for the number of bivariate tests performed (see Supplementary Data 8-9). Ventral and umbilical hernia shared the greatest number of local *r*_*g*_ correlations (n = 69; 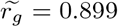) followed by inguinal and umbilical hernia (n = 60; 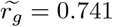 and ventral and inguinal hernia (n = 40; 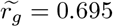) (see Figure 4A). These results contrast with genome-wide estimations of *r*_*g*_ where inguinal and femoral hernias were most correlated; however, they align with clinical classifications that specify inguinal hernia as a type of abdominal hernia alongside ventral and umbilical hernias.

**Figure 4:**
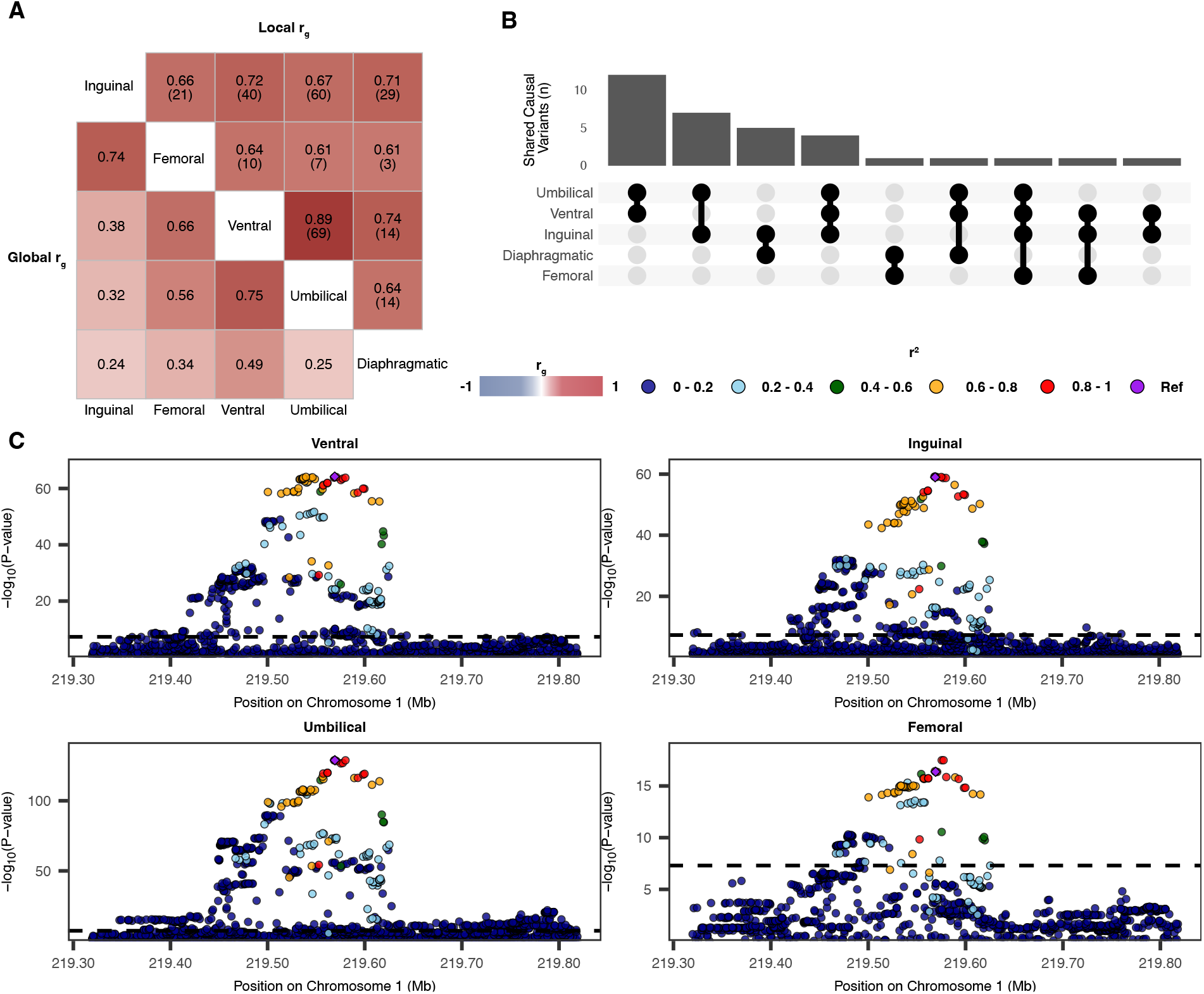
Genomic correlation and shared causal variants among hernia subtypes. **A** Heatmap demonstrating the global genomic correlation (lower triangle) and local genomic correlation (upper triangle) between hernia subtypes as estimated by LDSC and LAVA, respectively. The number in parentheses in the upper triangle indicates the number of significant pairwise loci after correcting for multiple testing (*P <* 0.05*/*961). **B** Upset plot demonstrating the number of loci that colocalized between each hernia subtype. **C** Regional locus plot for rs1415287 which was prioritized as a shared likely causal SNP between four hernia subtypes

Given the shared genome-wide significant loci and variants within credible sets, we also performed formal multi-trait colocalization analysis to determine whether hernia subtypes are influenced by the same causal signals at common genome-wide significant loci (see Methods). Colocalization analysis identified 33 loci with evidence of shared causal signals across hernia subtypes, including 30 loci with strong evidence for colocalization (PPFC > 0.7; see Supplementary Data 10-11). Colocalizing signals included 1:219569195:T:C located nearest *ZC3H11B* (PPFC = 0.9232 of which 38.6% is explained by the SNP; see Figure 4C), 17:56695877:T:G near *MFAP4* shared across ventral, diaphragmatic, and umbilical hernias (PPFC = 0.9953 of which 94.1% is explained by the SNP), and 2:55869757:A:G located near *EFEMP1* demonstrated strong evidence for colocalization across ventral, inguinal, and femoral hernias (PPFC = 0.9916 of which 99.9% is explained by the SNP). To identify biological pathways underlying shared hernia biology, we conducted gene set enrichment analysis of prioritized genes using the Reactome Pathway (2024) and GO Biological Process (2025) databases (see Supplementary Data 12). Reactome pathways demonstrated enrichment in “inactivation of anti-apoptotic BCL2 members” and “intrinsic pathways for apoptosis” (*BCL2* and *BMF*); extracellular matrix organization (*ADAMTS16, ADAMTS5, EFEMP1, COL24A1*, and *BMP7*); and elastic fiber formation (*EFEMP1* and *BMP7*; see Supplementary Figure 5A). GO Biological Processes demonstrated enrichment in mesonephric tubule, ureteric bud, and metanephric mesenchyme development (*WT1, OSR1, TCF21*, and *BMP7*; see Supplementary Figure 5B). Overall, these results underscore the importance of connective tissue biology in hernia development but also implicate embryologic development processes and pro-apoptotic signaling as important common factors for susceptibility to multiple hernia types.

### Genomic structural equation modeling reveals distinct genetic dimensions of hernia subtypes

Motivated by a high degree of genetic correlation and shared loci across traits, we modeled shared genetic liability across hernia subtypes with genomic structural equation modeling (Genomic-SEM). We first conducted exploratory factor analyses comparing one- and two-factor solutions under varimax and promax rotations (see Supplementary Data 13). Although a common factor accounted for 46.8% of the variance across subtypes, it exhibited only an adequate fit to the observed covariance (CFI = 0.84, SRMR = 0.11), suggesting residual heterogeneity beyond a single dimension. A two-factor solution yielded increased model fit (CFI = 0.92, SRMR = 0.10), suggesting a separation of hernia liability into two dimensions: a ventral/umbilical/diaphragmatic dimension and an inguinal/femoral dimension (see Figure 5A).

**Figure 5:**
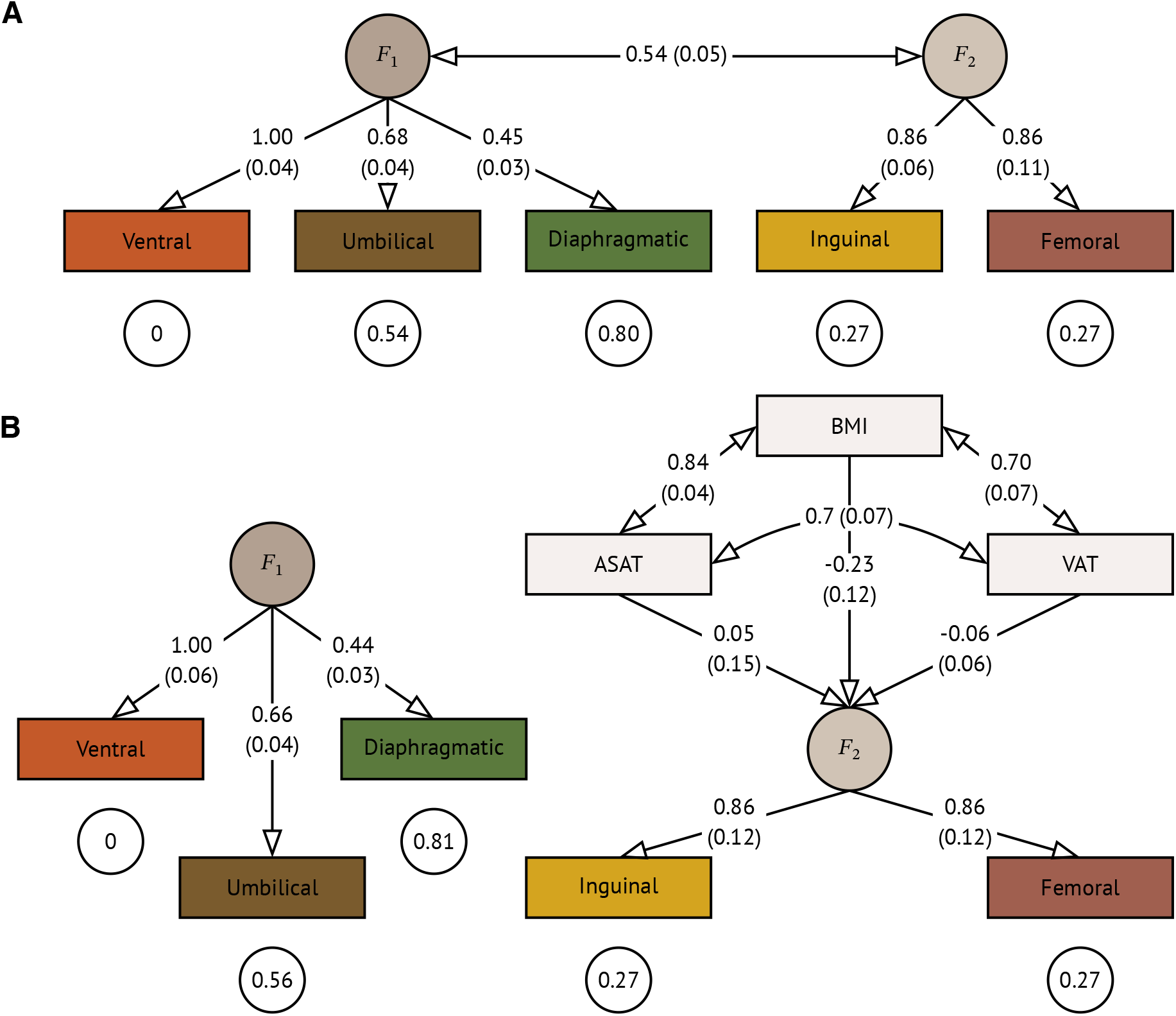
Structural equation diagrams of hernia genetic architecture. **A** Estimates from a two factor model with standard errors presented in parentheses and residual variances displayed within circles below each hernia type. Estimates represent the proportion of common variant liability accounted for by each latent factor. The two factor model suggested a separation of hernia liability into two dimensions: ventral/umbilical/diaphragmatic dimension and inguinal/femoral. **B** Estimates from a two factor model wherein *F*_2_ was regressed on body mass index, abdominal subcutaneous adipose tissue, and visceral adipose tissue.

Within this two-factor model, factor one was defined most strongly by ventral hernia and accounted for 46% and 20% of the common variant liability for umbilical and diaphragmatic hernia, respectively. In contrast, factor two accounted for the genetic liability for inguinal and femoral hernias evenly, explaining 73% of their common variant liability. In addition, the inter-factor correlations between the two hernia dimensions (*r*_*g*_ = 0.54, SE = 0.05) suggest roughly even contributions from general and specific genetic risk factors affecting liability for a specific hernia subtype.

Given the positive association of BMI with ventral and umbilical hernias and the negative association of BMI with inguinal and femoral hernias, we tested whether the shared genetic liability underlying these hernias, captured by factor two, was related to genetic liability for adiposity traits. Similar to the epidemiological literature, we observed negative genetic correlations between inguinal and femoral hernias and adiposity traits (see Supplementary Figure 8). Next, we applied the two-factor model and conducted a multiple regression, where factor two was regressed on BMI, visceral adipose tissue (VAT), and abdominal subcutaneous adipose tissue (ASAT). Interestingly, when controlling for their shared genetic variability, BMI exhibited a modest negative relationship with factor two (*β* = -0.23, SE = 0.12), whereas VAT and ASAT showed little evidence of independent association. These findings suggest that the shared genetic liability underlying inguinal and femoral hernia is not well explained by adiposity related genetic architecture.

### Mendelian randomization implicates causal phenotypes in hernia development

We performed Mendelian randomization analysis to identify putatively causal phenotypes in hernia formation by deriving high-confidence genetic instruments for known and suspected hernia risk factors from summary statistics of the most recent, largest available GWAS available on the GWAS Catalog (see Supplementary Data 14-16). Across all hernia subtypes, Mendelian randomization analyses demonstrated consistent evidence that genetically predicted anthropometric traits influence genetic liability to hernia (see Figure 6A). We found significant positive associations between BMI and ventral hernia (OR 1.69 per 1-SD increase; 95 % CI 1.62 to 1.77; FDR < 0.001), umbilical hernia (OR 1.54 per 1-SD increase; 95 % CI 1.47 to 1.62; FDR < 0.001), and diaphragmatic hernia (OR 1.18 per 1-SD increase; 95 % CI 1.12 to 1.24; FDR < 0.001). In contrast to other hernia subtypes, Mendelian randomization analyses revealed that higher BMI (OR 0.74 per 1-SD increase; 95% CI 0.69 to 0.90; FDR < 0.001), waist circumference (OR 0.77 per 1-SD increase; 95 % CI 0.67 to 0.86; FDR < 0.001), weight (OR 0.89 per 1-SD increase; 95 % CI 0.84 to 0.95; FDR < 0.001), and waist to hip ratio (OR 0.77 per 1-SD increase; 95 % CI 0.67 to 0.86; FDR < 0.001) were inversely associated with inguinal hernia risk. These findings recapitulate the well-described epidemiological paradox whereby higher BMI appears protective against inguinal hernia.

**Figure 6:**
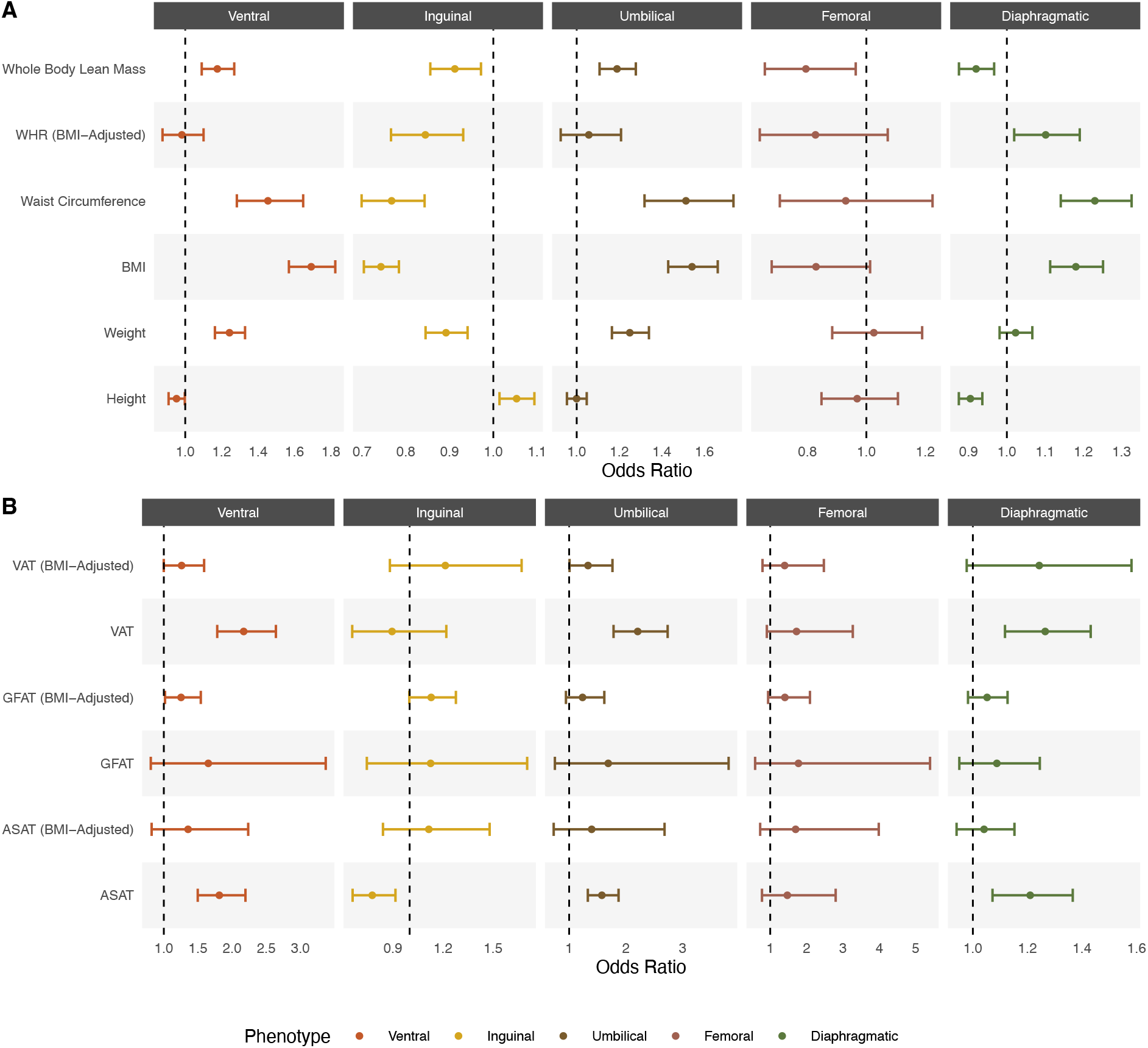
Mendelian randomization analysis across anthropometric and fat deposition traits.

Although BMI is a useful epidemiologic measure of overall adiposity for estimating hernia risk, it is an imprecise surrogate for the underlying anthropometric and biomechanical traits that may drive hernia pathogenesis. We therefore derived genetic instruments for fat distribution to evaluate how different fat deposition contributes to hernia risk. Consistent with the hypothesis that increased intra-abdominal pressure contributes to hernia formation, VAT was positively associated with ventral hernia, umbilical hernia, and diaphragmatic hernia (see Figure 6B and Supplementary Data 16). Similarly, ASAT showed positive associations with these phenotypes, potentially reflecting increased mechanical load and strain on abdominal wall structures. In contrast, inguinal hernia demonstrated inverse associations with both VAT and ASAT; however, these associations reversed in sign when using BMI-adjusted genetic instruments suggesting that VAT and ASAT increase inguinal hernia risk independent of BMI (see Figure 6B).

Outside of anthropometric measures, we tested whether selected phenotypes that could contribute to increased intra-abdominal pressure had a causal effect on hernia formation. We found a significant positive association between asthma and umbilical hernia (OR 1.11; 95% CI 1.04 to 1.18; FDR < 0.016), asthma and diaphragmatic hernia (OR 1.09; 95% CI 1.05 to 1.13; FDR < 0.001) and chronic obstructive pulmonary disease and diaphragmatic hernia (OR 1.17; 95% CI 1.09 to 1.25; FDR = 0.001). Lastly, we evaluated the role of smoking in hernia formation. We found a significant association between smoking initiation and ventral hernia (OR 1.83 per 1-SD increase; 95 % CI 1.70 to 1.96; FDR < 0.001), umbilical hernia (OR 1.31 per 1-SD increase; 95 % CI 1.18 to 1.44; FDR < 0.001), and diaphragmatic hernia (OR 1.39 per 1-SD increase; 95 % CI 1.28 to 1.49; FDR < 0.001). No significant associations between smoking and either inguinal hernia or femoral hernia were found.

### Proteome-wide and drug target Mendelian randomization identifies potential therapeutic targets for incisional hernia

Incisional hernias are a subset of ventral hernias that represent a challenging clinical phenotype due to poor risk prediction algorithms and high rates of recurrence after surgical treatment.^41;42^ We therefore performed an unbiased proteome-wide Mendelian randomization experiment using high confidence cis-acting genetic instruments for 1,881 proteins derived from a GWAS of proteomic data^43^ in order to identify potential therapeutic targets that would reduce risk of incisional hernia development following an abdominal surgery. To provide an added layer of evidence to the Mendelian randomization experiments, we performed an additional protein quantitative trait loci colocalization analysis as previously described.^44^ We identified 54 significant (FDR < 0.05) protein-trait associations across 49 distinct proteins and found seven phenotype-protein pairs with a high posterior probability for colocalization (PPFC > 0.7); see Supplementary Data 17-18).

Among these, MFAP4 was associated with inguinal hernia and diaphragmatic hernia. MFAP4 is an extracellular matrix protein that stabilizes elastic fibers and has been implicated in a range of connective tissue disorders including hernia, abdominal aortic aneurysm, and pelvic organ prolapse^13;45;46^. Under normal physiologic conditions, MFAP4 binds tropoelastin, Fibrillin-1, and Fibrillin-2 and promotes elastic fiber assembly^47^. However, in pathological states, it induces macrophage-driven inflammation and aberrant matrix metalloproteinase activity via integrin-*α*_*v*_*β*_3_/*α*_*v*_*β*_5_ mediated signaling and modulation of TGF-*β* activity, leading to maladaptive remodeling of the extracellular matrix (see Figure 7)^46;48–51^. Inhibition of this protein or its associated pathways may therefore be a promising target for a range of phenotypes including incisional hernia and abdominal aortic aneurysm. A monoclonal antibody against MFAP4 has recently shown efficacy in preclinical models of retinal neovascular disease^52^.

**Figure 7:**
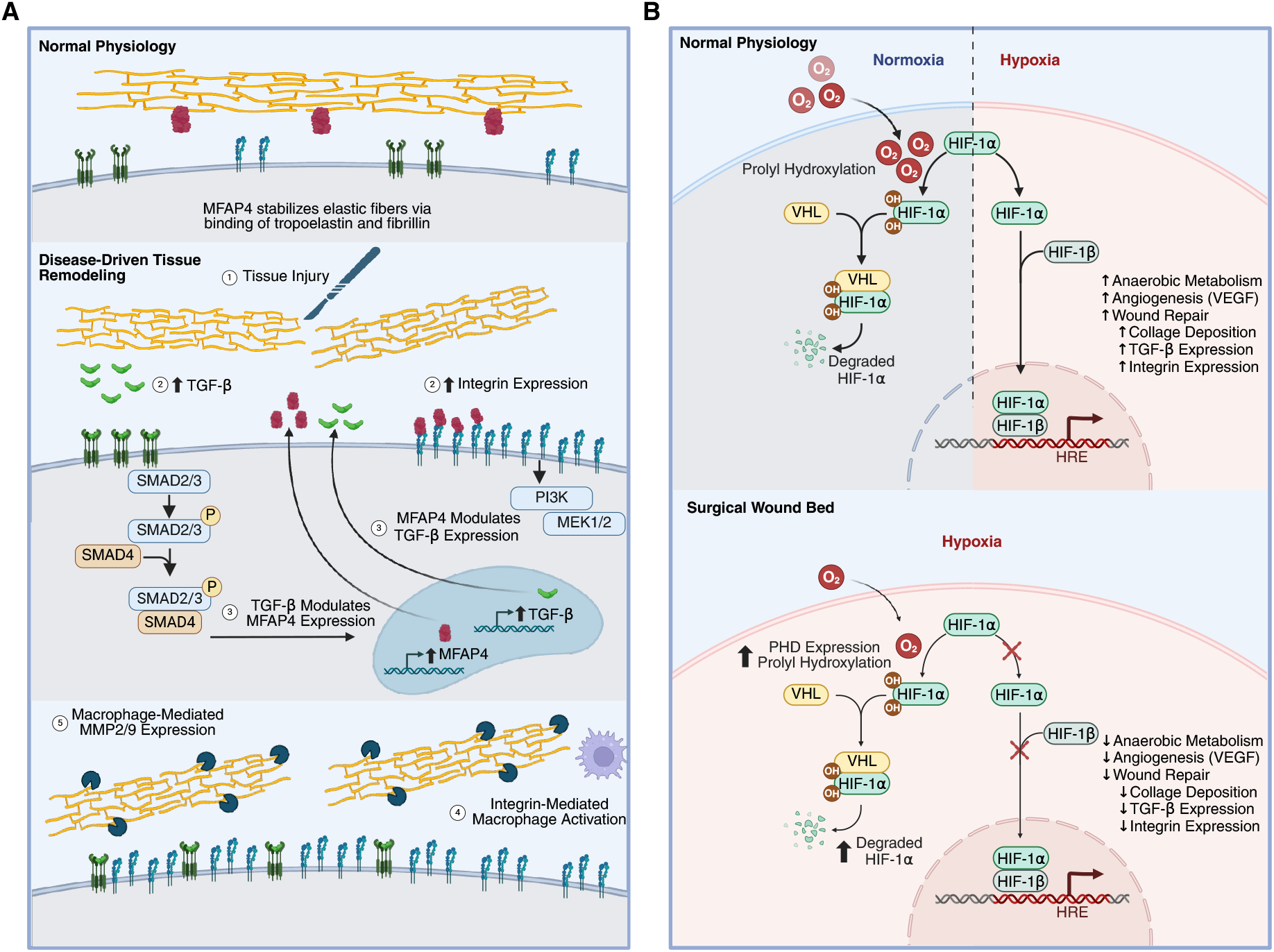
Proteome-wide and drug repurposing Mendelian randomization identify potential therapeutic targets for incisional hernia including (A) MFAP4 and (B) P4HTM. Created in BioRender.

We also performed drug repurposing Mendelian randomization analysis with previously described high confidence genetic instruments for currently approved or clinical-stage therapeutics to identify potential therapeutic targets for hernia formation.^53^ Across fibroblasts, subcutaneous adipose tissue, and visceral adipose tissue, we identified 51 unique drug targets, of which P4HTM was prioritized in all three tissues (see Supplementary Data 19). P4HTM is a transmembrane prolyl 4-hydroxylase that suppresses the activity of HIF-1*α* through post-translational hydroxylation of proline residues under normoxic conditions, marking it for proteasomal degradation.^54;55^ HIF-1*α* is a regulator of wound healing that induces gene expression changes which lead to increased angiogenesis, collagen deposition, TBG-*β* expression, and integrin expression.^54^ However, in pathological states such as diabetes, aging, and chronic ischemia, inappropriate P4HTM activity leads to suppression of HIF-1*α*, contributing to poor tissue healing and fascial weakening.^54;56^ Prolyl hydroxylase inhibitors have been approved for renal anemia and have shown preclinical efficacy in chronic and ischemic wound models, though none are currently approved for wound-healing indications and selectivity for P4HTM relative to other HIF-prolyl hydroxylases remains to be established.^54;57–59^

## Discussion

In this study, we present a comprehensive analysis of the overall genetic contributions to hernia development as well as the shared and distinct genetic architecture of hernia subtypes. We demonstrated that despite their anatomic diversity, hernia subtypes show a high degree of genomic similarity that converges on connective tissue and developmental biology pathways. Genomic-SEM specifically demonstrated a split between groin hernias and other abdominal wall hernias. Mendelian randomization implicated causal phenotypes, proteins, and therapeutic targets which overlap with other clinical phenotypes of interest including abdominal aortic aneurysms. Overall these findings raise important considerations for future studies of hernia genomics and identify features that could be used for future risk stratification and treatment.

We performed a multi-population genome-wide association meta analysis of multiple hernia phenotypes. Our multi-population analysis demonstrated improved power to detect novel hernia risk loci through both increased sample size and increased sample diversity. We replicated the majority of previously reported hernia risk loci, and report an additional 173 novel loci. This represents a three-fold increase in genomic discovery for hernia.^13^ In addition, our analyses expanded to include genetically diverse populations not previously assessed. Prior GWAS of hernia subtypes have largely been limited to individuals most similar to the 1000 Genomes European reference population.^13–15^ Here, we detect a novel locus located nearest *GRIK4* specific to individuals most genetically similar to the African reference population, underscoring the importance of multi-population studies in improving genomic discovery for complex traits. Future studies incorporating more data from diverse populations may enable further discovery of population-specific loci that improve our understanding of hernia biology and human genetics.

In analyzing the genomic architecture of multiple hernia traits, we found a high degree of overlap as evidenced by moderate to strong global and local *r*_*g*_ estimates, cross-trait shared causal variants identified through Bayesian colocalization analysis, and shared prioritized genes. Gene set enrichment analysis of prioritized genes reveals that biological pathways related to extracellular matrix formation and regulation, connective tissue biology, and embryologic development are key mediators of hernia risk. Despite these similarities, subtype specific differences do exist. For example in ventral hernia, variant-to-gene prioritization analysis demonstrated a missense variant located in *ITIH4* which is a type II acute-phase protein and extracellular matrix–stabilizing factor induced by IL-6 signaling in the hepatic acute-phase response to tissue injury.^60–63^ Dysregulation of this protein may therefore contribute to the development of incisional hernias which are labeled with ventral hernia diagnosis codes in contrast to the other hernia subtypes which are rarely the result of surgical procedures.

Our analysis identified two distinct latent genetic factors underlying hernia development. The first implicates inguinal and femoral hernias which are both forms of groin hernias. These hernia subtypes loaded more strongly onto their shared factor than the non-inguinofemoral subtypes did onto theirs. Although inguinal and femoral hernias are uniquely characterized by an inverse epidemiologic association with BMI, this latent factor appears genetically distinct from BMI and fat distribution traits, suggesting an independent biological mechanism. The second factor implicates ventral, umbilical, and diaphragmatic hernias, potentially consistent with a shared embryological origin as midline fusion defects. Diaphragmatic hernia exhibited residual genetic covariance beyond this factor, likely reflecting a greater congenital component and more distinctive developmental biology. Despite this two-factor structure, considerable residual genetic covariance exists between the factors, highlighting that shared biological pathways such as connective tissue biology underlie hernia susceptibility more broadly.

Through robust Mendelian randomization analysis, we also demonstrate that several phenotypes have putatuive causal associations. Of note, we found that visceral adipose tissue and abdominal subcutaneous adipose tissue have likely causal effects on hernia development while gluteofemoral adipose tissue does not. This finding highlights that BMI is correlated with the underlying physical features that contribute to hernia development. Future studies should explore the prognostic utility of imaging-derived measures of visceral adipose tissue and abdominal subcutaneous adipose tissue in hernia risk stratification, an approach which may offer more nuanced interpretation than BMI-based prediction. Our Mendelian randomization analysis also found that genetically predicted BMI, VAT, and ASAT were all associated with lower odds of inguinal hernia development. However, when using BMI-adjusted genetic instruments for VAT and ASAT, the associations for these features reversed sign: higher VAT and ASAT were associated with higher odds of inguinal hernia development. This pattern suggests that overall body mass may confound or mediate the observed associations and that increased BMI prevents diagnosis of inguinal hernia, leading to systematic ascertainment bias in observational data. Overall, this raises the possibility that BMI-related ascertainment or detection bias contributes to the observed inverse epidemiological relationship between BMI and inguinal hernia.

Our study must be interpreted in the context of its limitations. First, although we report the results of multi-population meta-analysis, our cohort is still predominately composed of individuals most genetically similar to the 1000 Genomes European reference population, and the population was not evenly distributed across hernia subtypes. For example, our analysis for femoral hernia was solely limited to the European population. Future analyses of hernia should strive to include more diverse populations to drive the development of our understanding of the common and distinct genomic architecture of hernia subtypes. Second, our effective sample size was not evenly distributed across hernia subtypes. There were approximately 50-times more inguinal hernia cases than femoral hernia cases, 5-times more inguinal hernia cases than ventral hernia cases, and 3-times more inguinal hernia cases than umbilical hernia cases. This imbalance prohibits us from identifying inguinal-hernia specific risk loci, as analyses in the other hernia subtypes may be underpowered to detect associations found in inguinal hernia. Third, the definition of hernia in each contributing cohort was based on diagnostic codes. Use of diagnostic coding data carries inherent risks of misclassification and ascertainment bias, and may further obscure subtype specific findings influencing the results of summary statistics. For example, inguinal hernia can be further subclassified as direct and indirect inguinal hernias, which are indistinguishable on physical exam but represent different pathology. Indirect inguinal hernias are congenital defects that result from failure of the processus vaginalis to close while direct inguinal hernias are believed to arise from increased intra-abdominal pressure. Though sharing a common anatomical presentation, these hernia subtypes may display different genomic architectures given their distinct pathophysiology. Finally, several X chromosome and sex-specific hernia-related risk loci have been reported. However, our summary data did not contain variants from the X chromosome and did not allow for sex-stratified analyses. Future studies of hernia could include sex stratified meta-analyses to identify and confirm sex specific loci.

## Methods

### Genome-wide association meta-analysis

We obtained GWAS summary statistics for five hernia subtypes–ventral, inguinal, umbilical, femoral, and diaphragmatic–from the All of Us Research Program,^16^ FinnGen,^17^ BioBank Japan,^18^ UK Biobank,^19^ and the VA Million Veteran Program^20^. Hernia case definitions were defined using biobank-specific phenotype definitions based on ICD-9, ICD-10, or PheCode categorizations. Prior to meta-analysis, we performed data harmonization and quality control with the following steps: (1) liftover from GrCh37 to GrCh38 was performed on summary statistics for the Pan-UK Biobank using the R package rtracklayer; GWASInspector was used to assess for test statistic inflation, skewness, and kurtosis;^64^ (3) allele harmonization was performed with the R package MungeSumstats using dbSNP v144 as a reference panel with the ‘bi_allelic_filter’ and ‘allele_flip_frq’ hyperparameters set to false;^65^ (4) LDSC was applied to any set of summary statistics with LDSC intercept > 1 to correct for genomic inflation using the Python package gwaslab^66^ with population-specific LD score files from Pan-UK Biobank;^67^ (5) a minor allele frequency filter was applied using the equation:

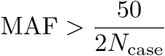

We then performed inverse-variance weighted fixed-effects meta-analysis with METAL for each hernia subtype within and between populations.^68^ In addition, we performed multi-population meta-regression using MR-MEGA (v0.2), allowing for four principal components.^69^ All summary statistics are presented on the GRCh38 assembly.

### Lead variant selection and statistical finemapping of hernia risk loci

We extracted independent lead variants from the meta-analyzed summary statistics using the Python package gwaslab.^66^ Loci were defined as the *±* 500 kb region flanking each lead variant (i.e., a 1-Mb window). Loci were classified as shared across hernia subtypes when these 1-Mb windows overlapped between lead variants from different subtypes, computed using the ‘genome_cluster’ function of the tidygenomics package in R (https://const-ae.github.io/tidygenomics/).

We performed statistical finemapping using approximate Bayes factors within the 1-Mb regions surrounding lead variants to derive 99% credible sets for locus–trait pairs. Specifically, we computed Bayes factors from GWAS effect estimates, standard errors, and sample size using the equation:

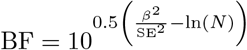

Posterior normalization were calculated using the equation:

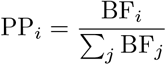

Variants were selected until the cumulative posterior probability reached 0.99.

### Multi-trait, expression quantitative trait loci, and protein quantitative trait loci colocalization analysis

We performed Bayesian colocalization analysis using HyPrColoc in R with default parameters.^70^ For multi-trait colocalization across hernia subtypes, we used multi-population meta-analyzed summary statistics within shared loci, defining locus boundaries as the minimum and maximum chromosomal positions across all overlapping lead variant windows. For eQTL colocalization, we tested the ±500 kb region surrounding each hernia lead variant. For pQTL colocalization, we used a ±250 kb window around each gene’s start and stop coordinates. A PPFC > 0.7 was considered high confidence for colocalization.

### Variant-to-gene prioritization and functional annotation

We performed variant-to-gene prioritization with an ensemble methodology comprised of annotating variants with their most severe consequence, calculating Polygenic Priority Scores,^28^ and performing eQTL colocalization analysis. Variant consequences were annotated using Ensembl VEP v113 executed via an offline Apptainer environment and the ‘–pick’ flag to select the most severe transcript consequence per variant.^27^ We applied PoPS using the default feature set and selected a PoPs prioritized gene if it had PoPs score in the 90th percentile for that hernia subtype. We performed eQTL colocalization analysis using HyPrColoc as above with fibroblast eQTL data from the eQTL Catalogue (Dataset Identifier: QTD000100).

### Heritability and cross-trait correlation estimation using linkage disequilibrium score regression

We estimated SNP-based heritability and cross-trait genetic correlations among hernia subtypes using LDSC. Liability-scale heritability was derived by converting observed-scale LDSC estimates using upper and lower bounds of population prevalence for each hernia subtype. We tested whether SNP-based heritability for hernia subtypes was enriched in genes with tissue- or cell type–specific expression using LDSC-SEG.^71^

### Local Analysis of Variance Association

We conducted local *r*_*g*_ analysis with LAVA v0.1.0.^40^ To identify significantly correlated local *r*_*g*_ between hernia subtypes, we first conducted univariate association testing at any genome-wide significant loci for all hernia subtypes requiring *P <* 0.05*/*243 before proceding with bivariate testing. This resulted in 961 bivariate tests, resulting in a P-value threshold of *P <* 0.05*/*961 to identify significant local *r*_*g*_ between hernia phenotypes. To perform the local *r*_*g*_ analysis conditional on body mass index, we also conducted univariate association testing at any genome-wide significant loci for hernia subtypes requiring *P <* 0.05*/*243. We accounted for sample overlap by providing LAVA with the estimated intercepts from bivariate LDSC calculated using the GenomicSEM R package.^72^

### Exploratory factor analysis and genomic structural equation modeling

To model shared genetic liability across hernia subtypes, we performed an exploratory and confirmatory factor analysis using the LDSC-derived genetic covariance structure from the GenomicSEM R package.^72^ We limited our analysis to European only meta-analyzed summary statistics. Exploratory factor analysis was performed using the R function factanal with common factor and two factor solutions under promax (correlated) varimax and promax (orthogonal) rotations. We performed confirmatory factor analysis in GenomicSEM; hernia subtypes were specified to load onto a factor if the exploratory factor analysis loadings were > 0.3. In the two factor model, we specified that the residual variances for ventral and inguinal hernia were > 0.001 to promote a parsimonious model fit.

### Mendelian randomization of anthropometric traits and other candidate causal hernia phenotypes

We performed two-sample Mendelian randomization to assess causal relationships between established hernia risk factors and hernia subtypes. We utilized inverse-variance weighted MR as the primary analysis when multiple SNPs were available for an exposure, and we utilized the Wald ratio approach when only one SNP was available for the exposure. We applied FDR correction to account for multiple testing, with q < 0.05 considered significant. Mendelian randomization analysis was performed with the R package TwoSampleMR.

### Proteome-wide Mendelian randomization

To identify circulating proteins causally associated with hernia phenotypes, we performed two-sample Mendelian randomization using genetic instruments for cis-acting protein quantitative trait loci (pQTL) for 1,881 plasma proteins.^43^ Outcome summary statistics derived from our meta-analysis. Mendelian randomization analysis and multiple testing correction was performed as described above. Additionally, we performed Bayesian colocalization analysis at each significant protein–trait locus as described above. Protein–trait pairs surviving both FDR-significant MR and high-confidence colocalization were prioritized as candidate therapeutic targets.

### Drug-repurposing Mendelain randomization

To identify approved or clinical-stage therapeutics whose targets may modulate hernia risk, we performed drug-target Mendelian Randomization using previously curated high-confidence genetic instruments for druggable genes.^53^ Tissue-specific eQTLs were obtained from GTEx v8 for three hernia-relevant tissues: cultured fibroblasts, subcutaneous adipose tissue, and visceral (omental) adipose tissue. Mendelian randomization analysis and multiple testing correction was performed as described above. Drug targets prioritized in all three tissues were considered the highest-confidence candidates for therapeutic repurposing.

### Gene set enrichment analyses

We performed gene set enrichment using Enrichr with the Reactome Pathways (2024) and GO Biological Processes (2025) to evaluate pathways implicated by prioritized/shared-variant genes.

## Declarations

### Data availability

Summary statistics from the genome-wide association study meta-analysis of ventral, inguinal, umbilical, femoral, diaphragmatic, and abdominal hernia will be deposited in the GWAS Catalog and made available upon publication.

### Code availability

All analyses were performed with publicly available code. Software packages and associated URLs are included as follows: R statistical software 4.3.2, https://www.R-project.org; MungeSumstats, https://github.com/Al-Murphy/MungeSumstats; LDSC 1.0.1, https://github.com/bulik/ldsc; gwaslab 3.6.3, https://github.com/Cloufield/gwaslab; METAL, https://genome.sph.umich.edu/wiki/METAL; MR-MEGA v0.2, https://genomics.ut.ee/en/tools; HyPrColoc 0.0.2 https://github.com/jrs95/hyprcoloc; MAGMA v1.10, https://ctg.cncr.nl/software/magma; PoPS https://github.com/FinucaneLab/pops; ENSEMBL VEP, https://github.com/Ensembl/ensembl-vep; TwoSampleMR 0.7.4 https://github.com/MRCIEU/TwoSampleMR; GenomicSEM https://github.com/GenomicSEM/GenomicSEM

## Acknowledgments

We gratefully acknowledge the participants of the UK Biobank, FinnGen, All of Us Research Program, BioBank Japan, and Million Veterans Program without whom this research would not have been possible. The AOU Research Program is supported by the National Institutes of Health, Office of the Director: Regional Medical Centers—1 OT2 OD026549, 1 OT2 OD026554, 1 OT2 OD026557, 1 OT2 OD026556, 1 OT2 OD026550, 1 OT2 OD 026552, 1 OT2 OD026553, 1 OT2 OD026548, 1 OT2 OD026551 and 1 OT2 OD026555; IAA #—AOD 16037; Federally Qualified Health Centers—HHSN 263201600085U; Data and Research Center—5 U2C OD023196; Biobank—1 U24 OD023121; The Participant Center—U24 OD023176; Participant Technology Systems Center—1 U24 OD023163; Communications and Engagement—3 OT2 OD023205 and 3 OT2 OD023206, and Community Partners—1 OT2 OD025277, 3 OT2 OD025315, 1 OT2 OD025337 and 1 OT2 OD025276. In addition, the AOU Research Program would not be possible without the partnership of its participants.

## Contributions

- AMP, JML, MGL, SMD, and HW designed the study.
- AMP, SY, JML, SAA, JD, RJ, GS contributed to data acquisition and analysis.
- AMP, JML, SMD, and HW drafted the paper.
- MGL, SMD, and HW supervised the work.
- All authors reviewed the paper and provided critical revisions.

## Ethics declarations

SY was supported by an Award from the American Heart Association (Award ID: 24POST1189614) and the VIVA Foundation. MGL receives grant funding from the Doris Duke Foundation (award 2023-2024) and US Department of Veterans Affairs Biomedical Research and Development (award IK2-BX006551), grant funding to the institution from MyOme, and consulting fees from BridgeBio. HW receives grant support from the National Cancer Institute of the National Institutes of Health (award #K08 CA270385). This work does not represent the views of the Department of Veterans Affairs or the US government.

## Supplementary Material

**Supplementary Figure 1:**
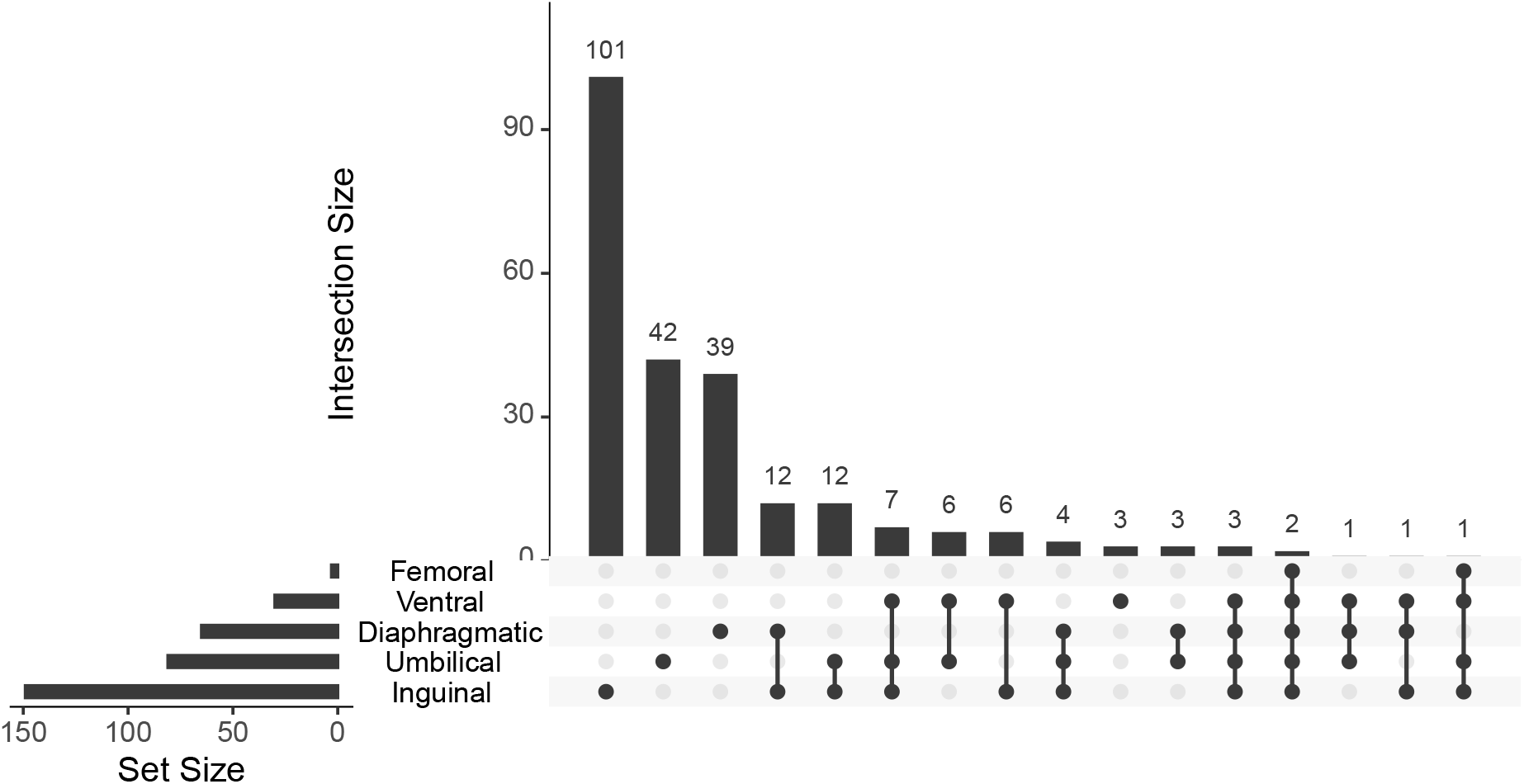
Upset plot demonstrating the number of shared genome-wide significant loci between each hernia subtype. Genome-wide significant loci were classified as shared between hernia subtypes if the 1 Mb region surrounding a lead variant overlapped with a 1 Mb region surrounding a different lead variant. Inguinal hernia shared the greatest number of loci with umbilical hernia (n = 12) and diaphragmatic hernia (n = 12). Two loci were shared across all five subtypes of hernia.

**Supplementary Figure 2:**
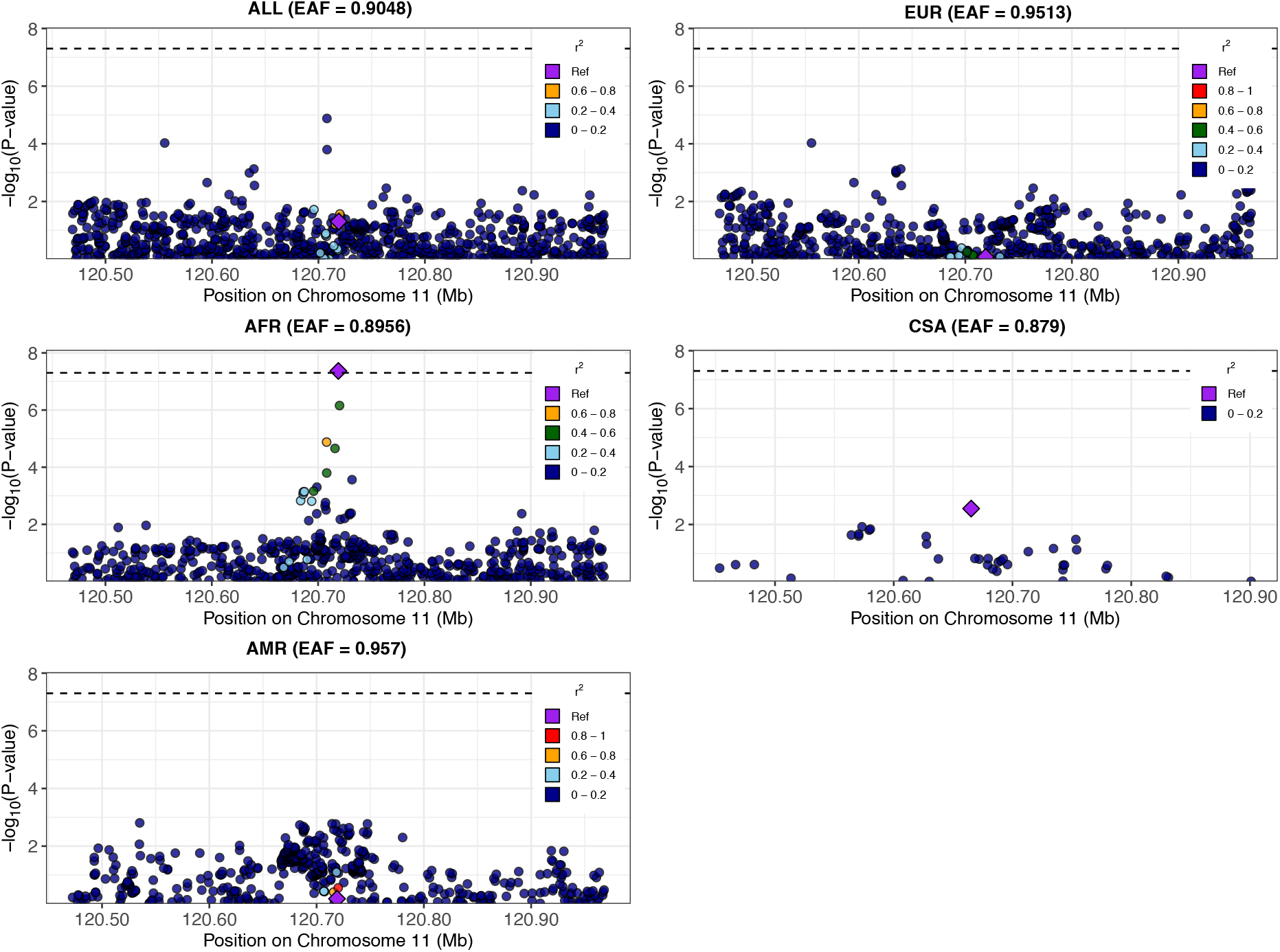
Regional association plots for locus located nearest *GRIK4* where a genome-wide significant variant was identified in individuals of African ancestry.

**Supplementary Figure 3:**
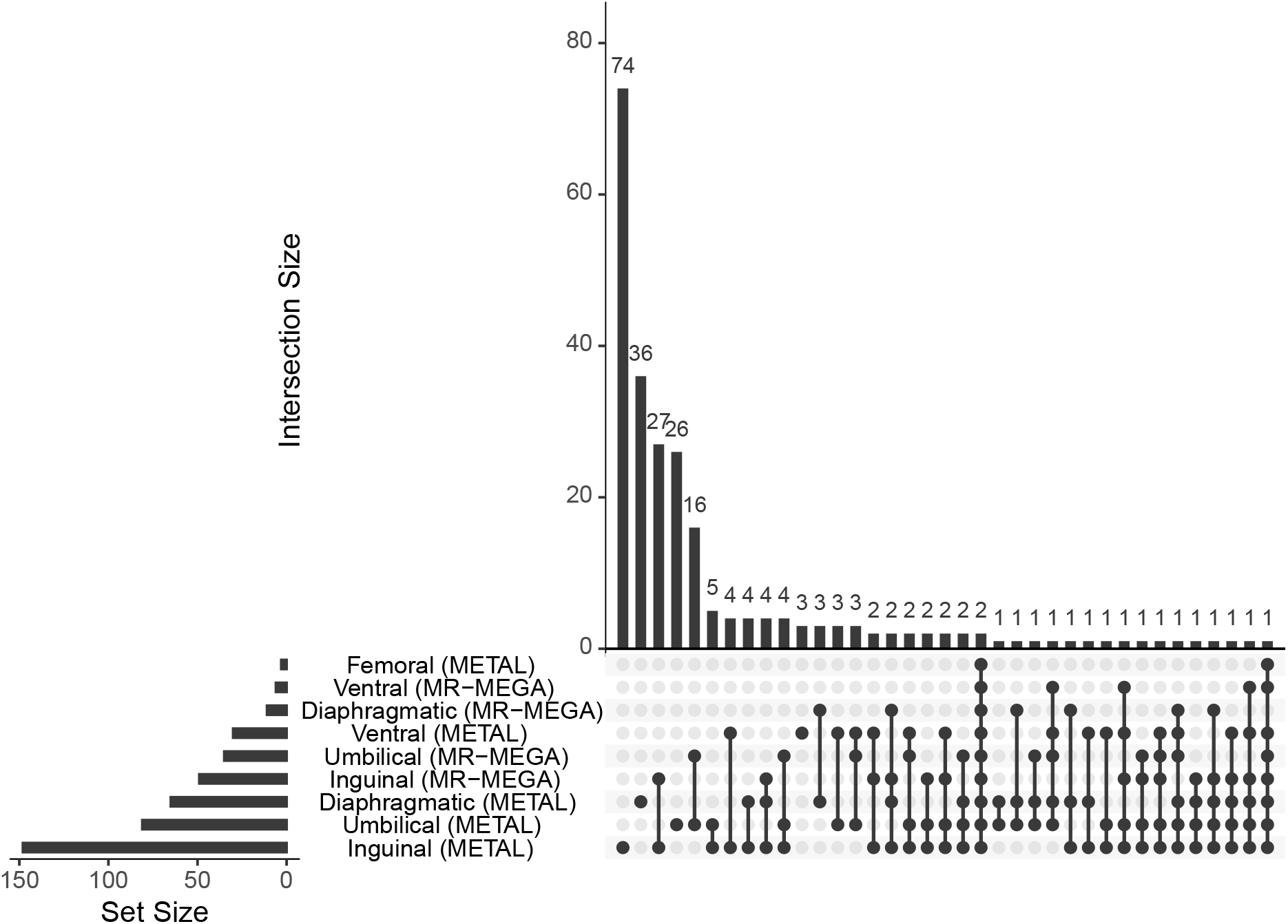
Upset plot demonstrating the number of shared genome-wide significant loci between each hernia subtype and meta-regression method. Genome-wide significant loci were classified as shared between hernia subtypes if the 1 Mb region surrounding a lead variant overlapped with a 1 Mb region surrounding a different lead variant. All genome-wide significant loci identified by MR-MEGA were also present in the METAL results; however, MR-MEGA did not identify any novel loci beyond the METAL based regression.

**Supplementary Figure 4:**
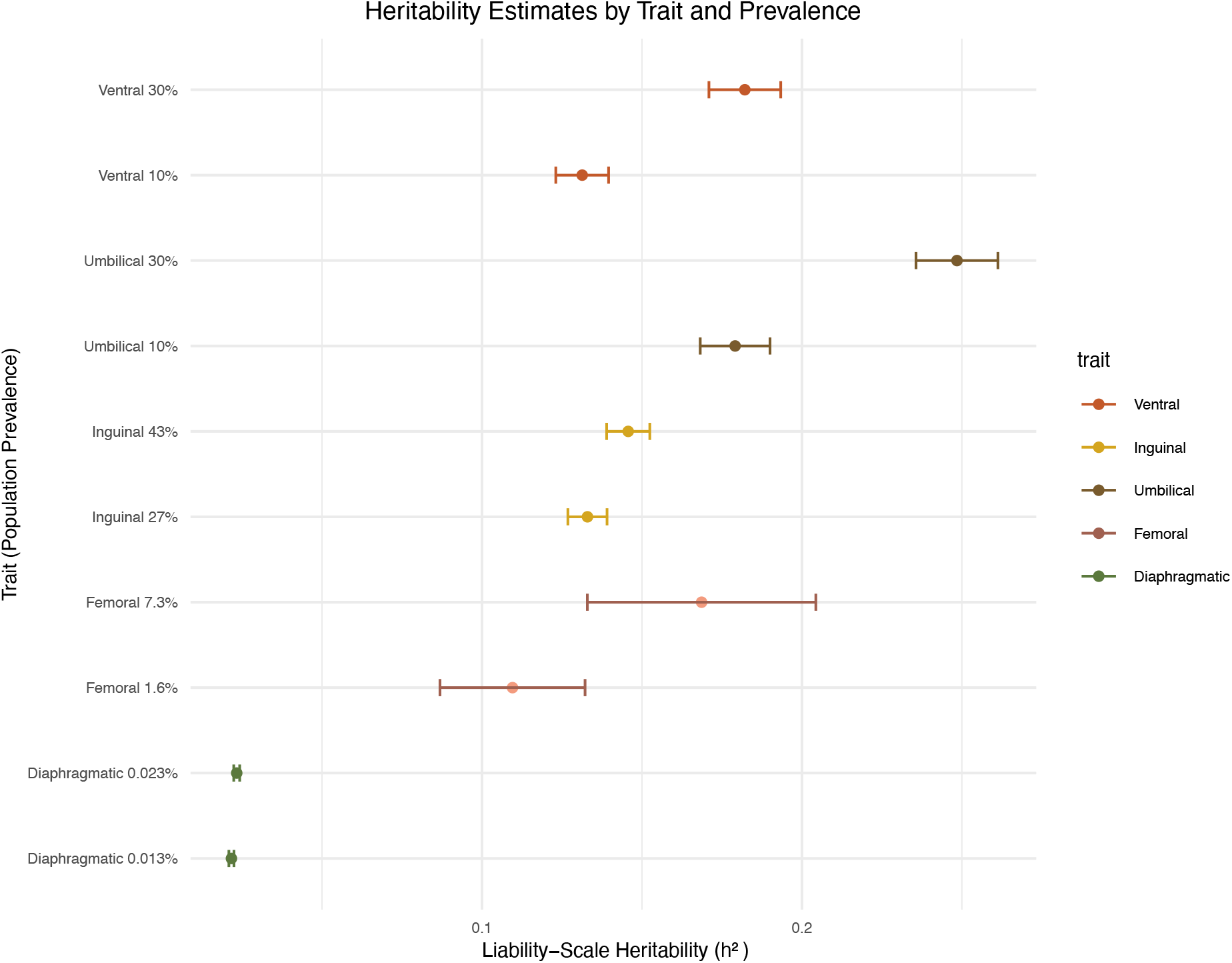
Heritability estimates for each subtype of hernia. Linkage disequilibrium score regression was applied to multi-population meta-regression summary statistics to estimate observed scaled heritability. Observed scale estimates were converted to liability scale estimates using upper and lower bounds of population prevalences for each hernia subtype.

**Supplementary Figure 5:**
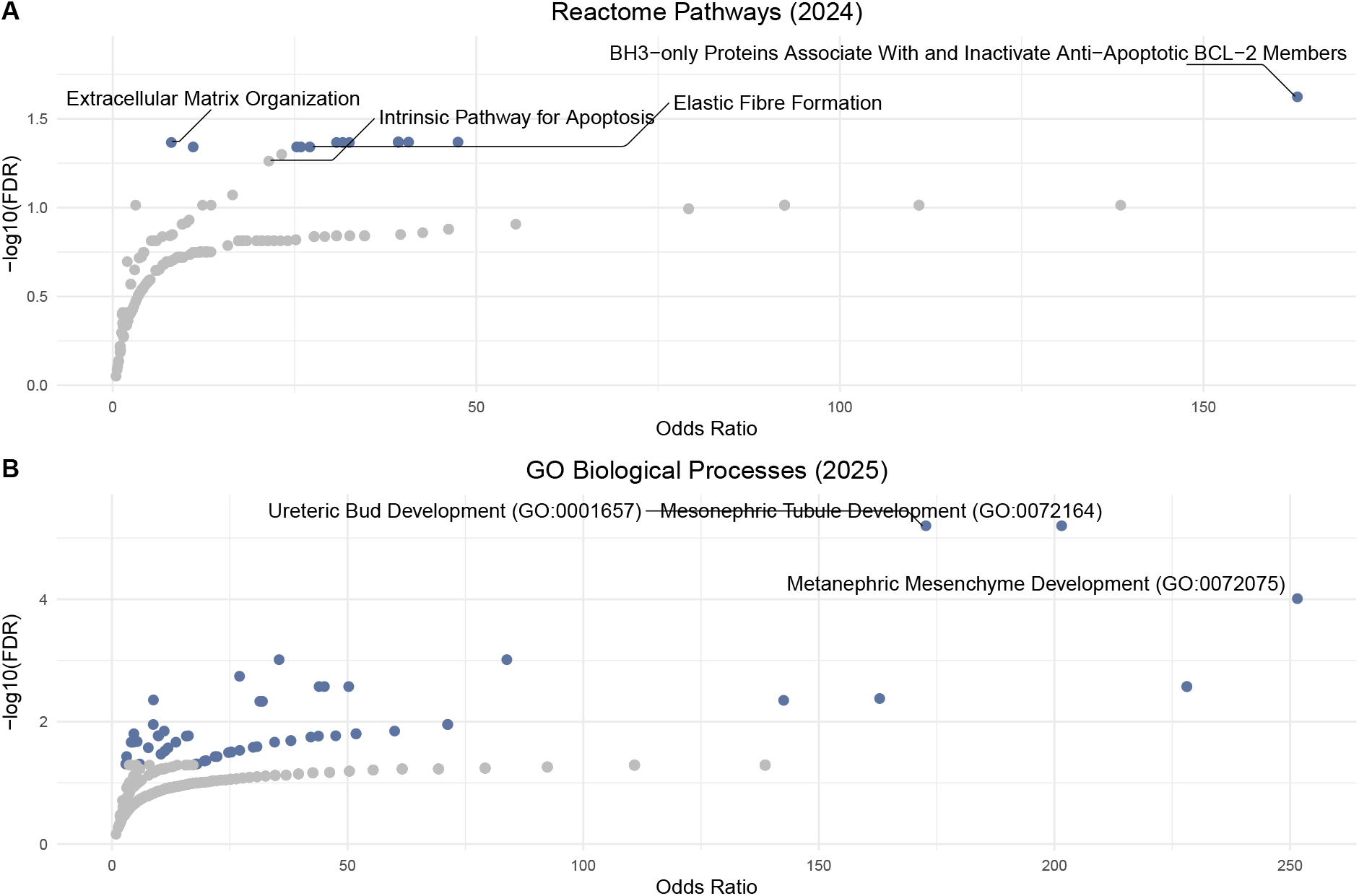
Most enriched gene ontologies in Reactome Pathways (2024) and GO Biological Processes for protein-coding genes nearest to genome-wide significant loci demonstrating evidence for colocalization across hernia subtypes.

**Supplementary Figure 6:**
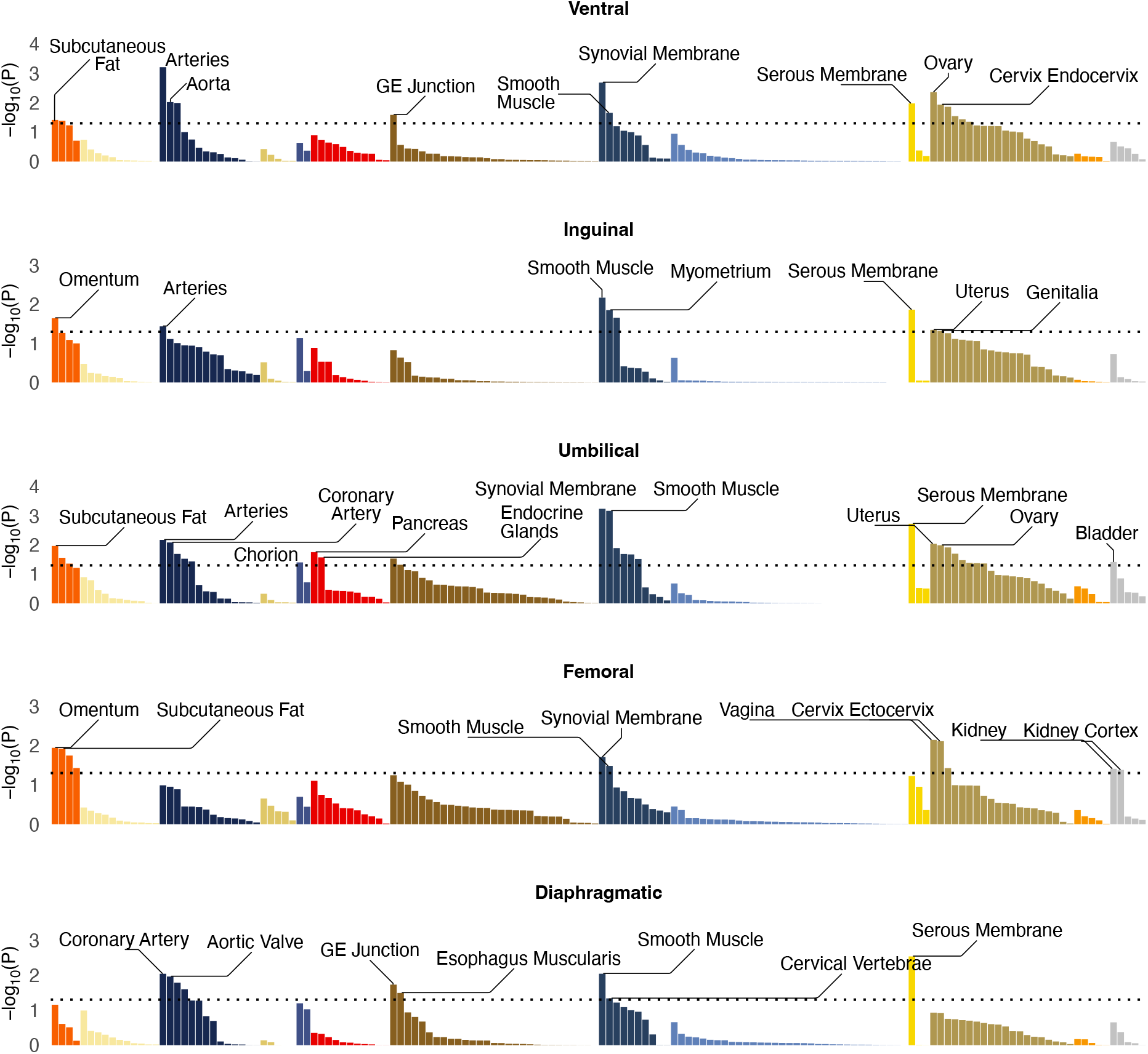
Tissue enrichment analysis of hernia subtypes using LDSC-SEG. Enrichment for tissue-specific differentially expressed genes, showing strongest signals in adipose and vascular tissues. The dashed black line indicates the nominal significance threshold (P = 0.05). Tissue groups are color-coded by organ system.

**Supplementary Figure 7:**
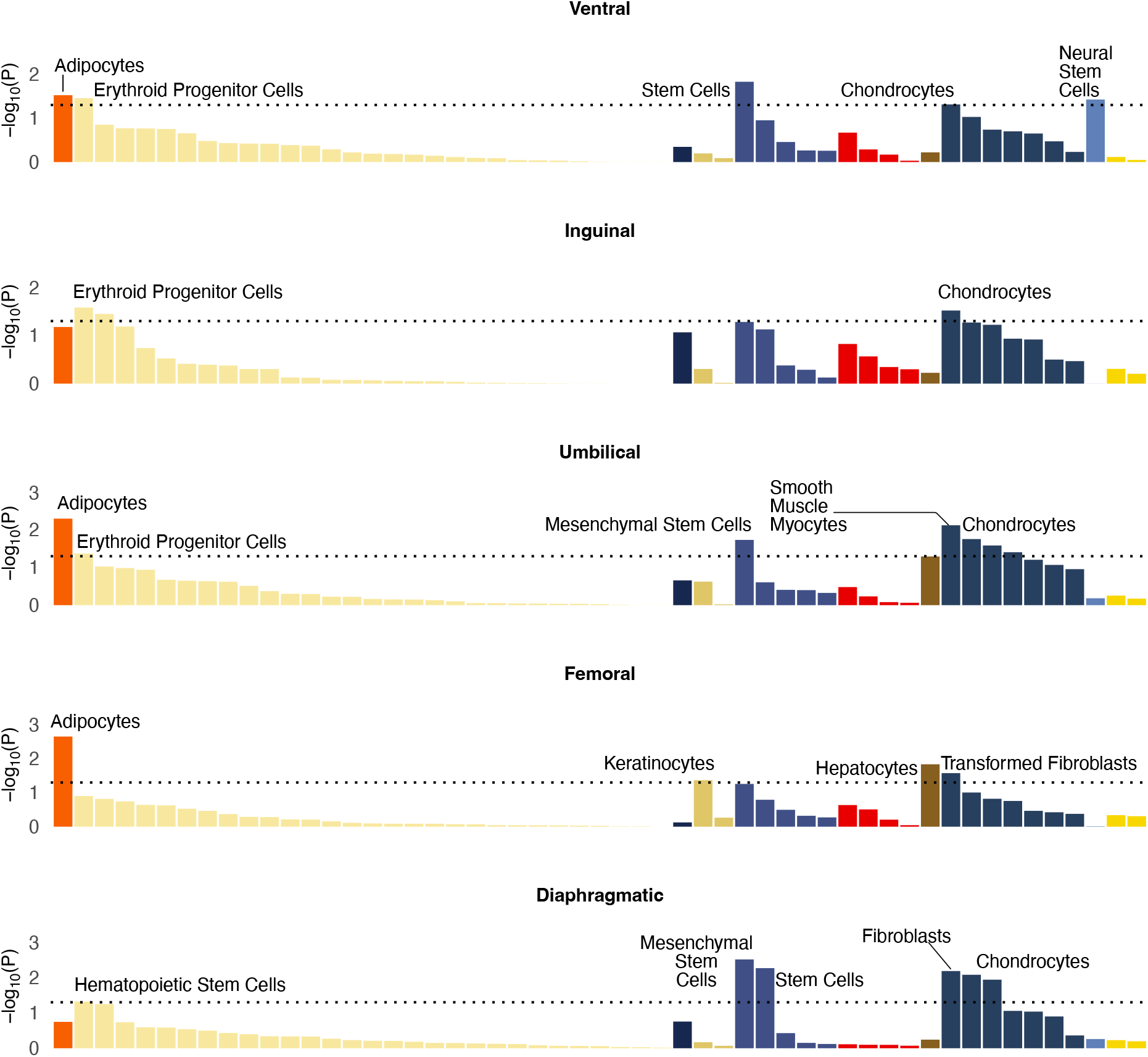
Cell type enrichment analysis of hernia subtypes using LDSC-SEG. Enrichment for cell-specific differentially expressed genes, showing strongest signals in mesenchymal stem cells, chondrocytes, adipocytes, and fibroblasts. The dashed black line indicates the nominal significance threshold (P = 0.05). Tissue groups are color-coded by organ system.

**Supplementary Figure 8:**
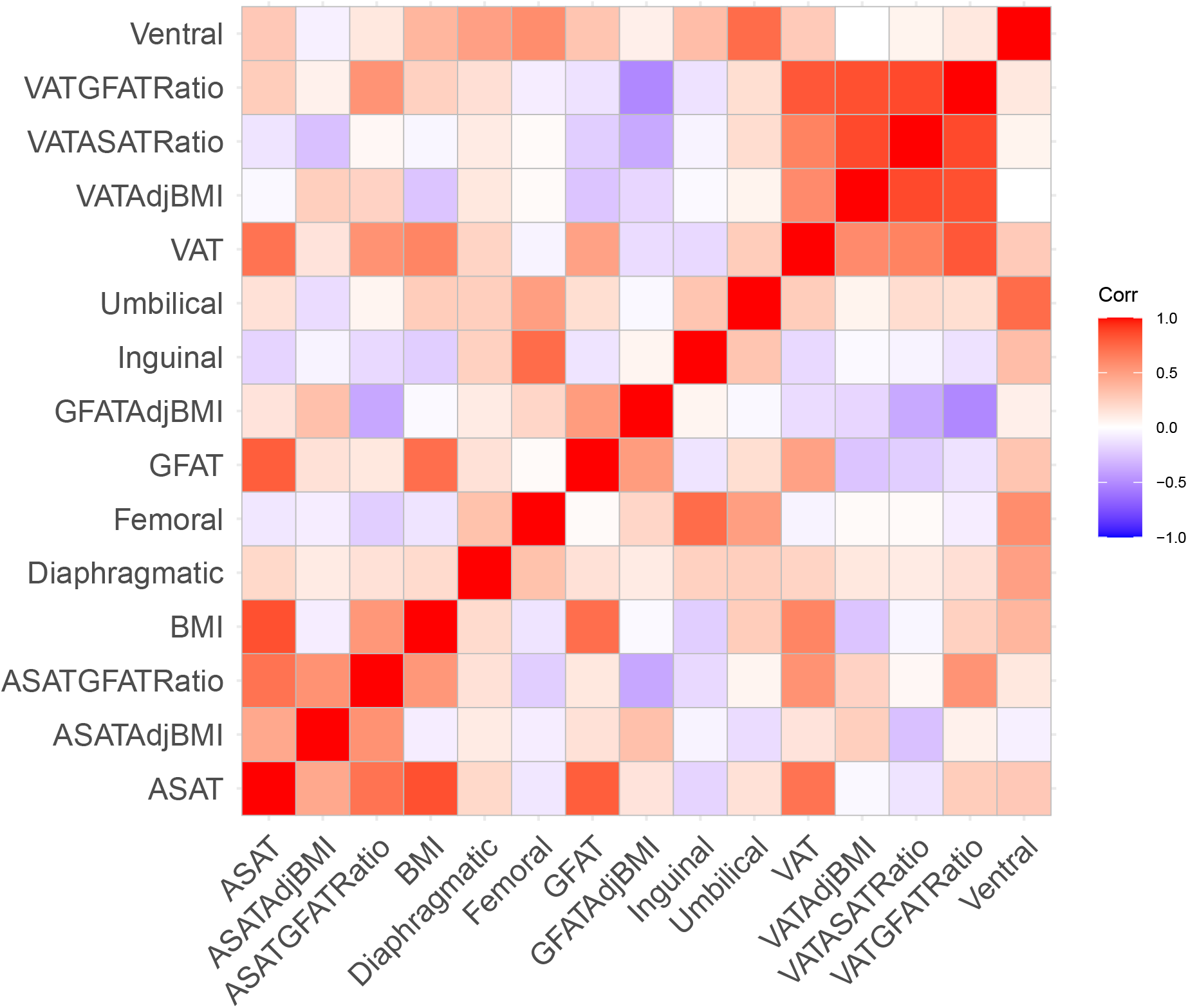
Genomic correlation between hernia subtypes and anthropometric traits

## Description of Supplementary Data

1. Number of participants with and without hernia subtypes, stratified by biobank and most similar 1000 Genomes reference population.
2. Lambda GC estimates and number of genome-wide significant variants for the meta-analyses, stratified by hernia subtype and population.
3. Lead variants identified in the multi-population meta analysis for each hernia subtype.
4. Credible sets containing less than ten variants for each hernia subtype.
5. Per-locus summary of variant-to-gene prioritization.
6. Gene set enrichment analysis of prioritized hernia effector genes using GO Biological Processes (2025).
7. Gene set enrichment analysis of prioritized hernia effector genes using the Reactome Pathways (2024).
8. Results of LAVA univariate analysis.
9. Results of LAVA bivariate analysis.
10. Results of Bayesian multi-trait colocalization analysis.
11. Statistical finemapping for causal variants at loci identified in colocalization analysis.
12. Gene set enrichment analysis of prioritized genes at loci identified in colocalization analysis
13. Results from exploratory factor analysis of hernia subtypes.
14. Summary details of Mendelian Randomization instruments.
15. Mendelian randomization instruments.
16. Mendelian randomization results.
17. Results of proteome-wide Mendelian randomization.
18. Results of Bayesian pQTL colocalization analysis.
19. Results of drug repurposing Mendelian randomization.

## References

[1] Ma, Q. et al. The global, regional, and national burden and its trends of inguinal, femoral, and abdominal hernia from 1990 to 2019: Findings from the 2019 Global Burden of Disease Study - a cross-sectional study. International Journal of Surgery 109, 333–342 (2023).

[2] Wang, F., Ma, B., Ma, Q. & Liu, X. Global, regional, and national burden of inguinal, femoral, and abdominal hernias: A systematic analysis of prevalence, incidence, deaths, and DALYs with projections to 2030. International Journal of Surgery 110, 1951–1967 (2024).

[3] Dai, F. et al. Spatiotemporal trends in hernia disease burden and health workforce correlations in aging populations: A global analysis with projections to 2050. BMC Gastroenterology 25, 296 (2025).

[4] Liu, X., Ma, B. & Ma, Q. Global pattern, trend and cross-country inequalities of inguinal, femoral, and abdominal hernia among individuals aged 60 and above from 1990 to 2021 and projections until 2040: A population-based study. Surgical Endoscopy 39, 4335–4344 (2025).

[5] Lau, B., Kim, H., Haigh, P. I. & Tejirian, T. Obesity increases the odds of acquiring and incarcerating noninguinal abdominal wall hernias. The American Surgeon 78, 1118–1121 (2012).

[6] Zelicha, H., Bell, D. S., Chen, D., Chen, Y. & Livingston, E. H. Obesity and abdominal hernia in ambulatory patients, 2018-2023. Hernia: The Journal of Hernias and Abdominal Wall Surgery 28, 1317–1324 (2024).

[7] Zöller, B., Ji, J., Sundquist, J. & Sundquist, K. Shared and nonshared familial susceptibility to surgically treated inguinal hernia, femoral hernia, incisional hernia, epigastric hernia, and umbilical hernia. Journal of the American College of Surgeons 217, 289–299.e1 (2013).

[8] Burcharth, J., Pommergaard, H. C. & Rosenberg, J. The inheritance of groin hernia: A systematic review. Hernia: The Journal of Hernias and Abdominal Wall Surgery 17, 183–189 (2013).

[9] Öberg, S., Sæter, A. H. & Rosenberg, J. The inheritance of groin hernias: An updated systematic review with meta-analyses. Hernia: The Journal of Hernias and Abdominal Wall Surgery 27, 1339–1350 (2023).

[10] Lau, H., Fang, C., Yuen, W. K. & Patil, N. G. Risk factors for inguinal hernia in adult males: A case-control study. Surgery 141, 262–266 (2007).

[11] Liem, M. S., van der Graaf, Y., Beemer, F. A. & van Vroonhoven, T. J. Increased risk for inguinal hernia in patients with Ehlers-Danlos syndrome. Surgery 122, 114–115 (1997).

[12] Chang, H.-H., Juan, Y.-S., Li, C.-C., Lee, H.-Y. & Chen, J.-H. Congenital collagenopathies increased the risk of inguinal hernia developing and repair: Analysis from a nationwide population-based cohort study. Scientific Reports 12, 2360 (2022).

[13] Fadista, J. et al. Comprehensive genome-wide association study of different forms of hernia identifies more than 80 associated loci. Nature Communications 13, 3200 (2022).

[14] Ahmed, W. U.-R. et al. Shared genetic architecture of hernias: A genome-wide association study with multivariable meta-analysis of multiple hernia phenotypes. PloS One 17, e0272261 (2022).

[15] Wei, J. et al. Identification of fifty-seven novel loci for abdominal wall hernia development and their biological and clinical implications: Results from the UK Biobank. Hernia: The Journal of Hernias and Abdominal Wall Surgery 26, 335–348 (2022).

[16] The All of Us Research Program Genomics Investigators et al. Genomic data in the All of Us Research Program. Nature 627, 340–346 (2024).

[17] Kurki, M. I. et al. FinnGen provides genetic insights from a well-phenotyped isolated population. Nature 613, 508–518 (2023).

[18] Nagai, A. et al. Overview of the BioBank Japan Project: Study design and profile. Journal of Epidemiology 27, S2–S8 (2017).

[19] Sudlow, C. et al. UK biobank: An open access resource for identifying the causes of a wide range of complex diseases of middle and old age. PLoS Medicine 12, e1001779 (2015).

[20] Gaziano, J. M. et al. Million Veteran Program: A mega-biobank to study genetic influences on health and disease. Journal of Clinical Epidemiology 70, 214–223 (2016).

[21] The 1000 Genomes Project Consortium et al. A global reference for human genetic variation. Nature 526, 68–74 (2015).

[22] Baldarelli, R. M. et al. Mouse Genome Informatics: An integrated knowledgebase system for the laboratory mouse. Genetics 227, iyae031 (2024).

[23] Di Paolo, G. et al. Recruitment and regulation of phosphatidylinositol phosphate kinase type 1γ by the FERM domain of talin. Nature 420, 85–89 (2002).

[24] Qi, L. et al. Talin2-mediated traction force drives matrix degradation and cell invasion. Journal of Cell Science 129, 3661–3674 (2016).

[25] Golubkov, V. S. et al. The Wnt/planar cell polarity protein-tyrosine kinase-7 (PTK7) is a highly efficient proteolytic target of membrane type-1 matrix metalloproteinase: Implications in cancer and embryogenesis. The Journal of Biological Chemistry 285, 35740–35749 (2010).

[26] Sato, H. et al. A matrix metalloproteinase expressed on the surface of invasive tumour cells. Nature 370, 61–65 (1994).

[27] McLaren, W. et al. The Ensembl Variant Effect Predictor. Genome Biology 17, 122 (2016).

[28] Weeks, E. M. et al. Leveraging polygenic enrichments of gene features to predict genes underlying complex traits and diseases. Nature Genetics 55, 1267–1276 (2023).

[29] Zhu, Y. et al. Phosphorylation of Tara by Plk1 is essential for faithful chromosome segregation in mitosis. Experimental Cell Research 318, 2344–2352 (2012).

[30] Lan, J. et al. The 68-kDa Telomeric Repeat Binding Factor 1 (TRF1)-associated Protein (TAP68) Interacts with and Recruits TRF1 to the Spindle Pole during Mitosis. Journal of Biological Chemistry 289, 14145–14156 (2014).

[31] Yu, J. et al. The E3 ubiquitin ligase HECTD3 regulates ubiquitination and degradation of Tara. Biochemical and Biophysical Research Communications 367, 805–812 (2008).

[32] Bradshaw, N. J. et al. An unpredicted aggregation-critical region of the actin-polymerizing protein TRIOBP-1/Tara, determined by elucidation of its domain structure. Journal of Biological Chemistry 292, 9583–9598 (2017).

[33] Tucker, E. J. et al. Mutations in the UQCC1-interacting protein, UQCC2, cause human complex III deficiency associated with perturbed cytochrome b protein expression. PLoS Genetics 9, e1004034 (2013).

[34] Kearsey, J., Petit, S., De Oliveira, C. & Schweighoffer, F. A novel four transmembrane spanning protein, CLP24. A hypoxically regulated cell junction protein. European Journal of Biochemistry 271, 2584–2592 (2004).

[35] Bai, S. W. et al. Identification and characterization of a set of conserved and new regulators of cytoskeletal organization, cell morphology and migration. BMC Biology 9, 54 (2011).

[36] Wong, G. W. et al. Molecular, biochemical and functional characterizations of C1q/TNF family members: Adipose-tissue-selective expression patterns, regulation by PPAR-gamma agonist, cysteine-mediated oligomerizations, combinatorial associations and metabolic functions. The Biochemical Journal 416, 161–177 (2008).

[37] Li, K. et al. CTRP7 Is a Biomarker Related to Insulin Resistance and Oxidative Stress: Cross-Sectional and Intervention Studies In Vivo and In Vitro. Oxidative Medicine and Cellular Longevity 2022, 6877609 (2022).

[38] Petersen, P. S. et al. CTRP7 deletion attenuates obesity-linked glucose intolerance, adipose tissue inflammation, and hepatic stress. American Journal of Physiology. Endocrinology and Metabolism 312, E309–E325 (2017).

[39] Hu, W. et al. Circulating CTRP7 Is a Potential Predictor for Metabolic Syndrome. Frontiers in Endocrinology 12, 774309 (2021).

[40] Werme, J., van der Sluis, S., Posthuma, D. & de Leeuw, C. A. An integrated framework for local genetic correlation analysis. Nature Genetics 54, 274–282 (2022).

[41] Prevention of incisional hernia with prophylactic onlay and sublay mesh reinforcement vs. primary suture only in midline laparotomies (PRIMA): long-term outcomes of a multicentre, double-blind, randomised controlled trial. The Lancet Regional Health - Europe 36, 100787 (2024).

[42] Thankam, F. G., Palanikumar, G., Fitzgibbons, R. J. & Agrawal, D. K. Molecular Mechanisms and Potential Therapeutic Targets in Incisional Hernia. The Journal of Surgical Research 236, 134–143 (2019).

[43] Pietzner, M. et al. Mapping the proteo-genomic convergence of human diseases. Science 374 (2021).

[44] Levin, M. G. et al. Genetics of varicose veins reveals polygenic architecture and genetic overlap with arterial and venous disease. Nature Cardiovascular Research 2, 44–57 (2023).

[45] Xie, Z., Feng, Y., He, Y., Lin, Y. & Wang, X. Identification of potential drug targets for pelvic organ prolapse using a proteome-wide Mendelian randomization approach. Scientific Reports 15, 8291 (2025).

[46] Kanaan, R., Medlej-Hashim, M., Jounblat, R., Pilecki, B. & Sorensen, G. L. Microfibrillar-associated protein 4 in health and disease. Matrix Biology: Journal of the International Society for Matrix Biology 111, 1–25 (2022).

[47] Pilecki, B. et al. Characterization of Microfibrillar-associated Protein 4 (MFAP4) as a Tropoelastin- and Fibrillin-binding Protein Involved in Elastic Fiber Formation. Journal of Biological Chemistry 291, 1103–1114 (2016).

[48] Pan, Z. et al. MFAP4 deficiency alleviates renal fibrosis through inhibition of NF-κB and TGF-β/Smad signaling pathways. The FASEB Journal 34, 14250–14263 (2020).

[49] Pilecki, B. et al. MFAP4 Deficiency Attenuates Angiotensin II-Induced Abdominal Aortic Aneurysm Formation Through Regulation of Macrophage Infiltration and Activity. Frontiers in Cardiovascular Medicine 8, 764337 (2021).

[50] Wang, H.-B. et al. Deletion of Microfibrillar-Associated Protein 4 Attenuates Left Ventricular Remodeling and Dysfunction in Heart Failure. Journal of the American Heart Association 9, e015307 (2020).

[51] Wang, H., Liu, M., Wang, X., Shuai, W. & Fu, H. MFAP4 deletion attenuates the progression of angiotensin II-induced atrial fibrosis and atrial fibrillation. Europace 24, 340–347 (2022).

[52] Schlosser, A. et al. Pharmacological blocking of microfibrillar-associated protein 4 reduces retinal neoangiogenesis and vascular leakage. Molecular Therapy: The Journal of the American Society of Gene Therapy 33, 1048–1072 (2025).

[53] Gaziano, L. et al. Actionable druggable genome-wide Mendelian randomization identifies repurposing opportunities for COVID-19. Nature Medicine 27, 668–676 (2021).

[54] Hong, W. X. et al. The Role of Hypoxia-Inducible Factor in Wound Healing. Advances in Wound Care 3, 390–399 (2014).

[55] Koivunen, P. et al. An Endoplasmic Reticulum Transmembrane Prolyl 4-Hydroxylase Is Induced by Hypoxia and Acts on Hypoxia-inducible Factor-α. Journal of Biological Chemistry 282, 30544–30552 (2007).

[56] Chang, E. I. et al. Age decreases endothelial progenitor cell recruitment through decreases in hypoxia-inducible factor 1alpha stabilization during ischemia. Circulation 116, 2818–2829 (2007).

[57] Zhu, Y., Wang, Y., Jia, Y., Xu, J. & Chai, Y. Roxadustat promotes angiogenesis through HIF-1α/VEGF/VEGFR2 signaling and accelerates cutaneous wound healing in diabetic rats. Wound Repair and Regeneration 27, 324–334 (2019).

[58] Lokmic, Z., Musyoka, J., Hewitson, T. D. & Darby, I. A. Hypoxia and hypoxia signaling in tissue repair and fibrosis. International Review of Cell and Molecular Biology 296, 139–185 (2012).

[59] Botusan, I. R. et al. Stabilization of HIF-1alpha is critical to improve wound healing in diabetic mice. Proceedings of the National Academy of Sciences of the United States of America 105, 19426–19431 (2008).

[60] Chen, X.-Y. et al. Alveolar epithelial inter-alpha-trypsin inhibitor heavy chain 4 deficiency associated with senescence-regulated apoptosis by air pollution. Environmental Pollution 278, 116863 (2021).

[61] Kashyap, R. S. et al. Inter-alpha-trypsin inhibitor heavy chain 4 is a novel marker of acute ischemic stroke. Clinica Chimica Acta; International Journal of Clinical Chemistry 402, 160–163 (2009).

[62] Cai, H. et al. Quantitative proteomics analysis of radiation-induced rectal injury in rectal cancer patients undergoing radiotherapy. Journal of Proteomics 319, 105485 (2025).

[63] Li, J. & Liu, G. Lentiviral Injection of Inter-α-Trypsin Inhibitor Heavy Chain 4 Promotes Female Spinal Cord Injury Mice Recuperation by Diminishing Peripheral and Central Inflammation. Inflammation 48, 2387–2394 (2024).

[64] Ani, A., Van Der Most, P. J., Snieder, H., Vaez, A. & Nolte, I. M. GWASinspector: Comprehensive quality control of genome-wide association study results. Bioinformatics 37, 129–130 (2021).

[65] Murphy, A. E., Schilder, B. M. & Skene, N. G. MungeSumstats: A Bioconductor package for the standardization and quality control of many GWAS summary statistics. Bioinformatics 37, 4593–4596 (2021).

[66] He, Y., Koido, M., Shimmori, Y. & Kamatani, Y. GWASLab: A Python package for processing and visualizing GWAS summary statistics (2023).

[67] Karczewski, K. J. et al. Pan-UK Biobank GWAS improves discovery, analysis of genetic architecture, and resolution into ancestry-enriched effects. Nature Genetics 57 (2025).

[68] Willer, C. J., Li, Y. & Abecasis, G. R. METAL: Fast and efficient meta-analysis of genomewide association scans. Bioinformatics 26, 2190–2191 (2010).

[69] Mägi, R. et al. Trans-ethnic meta-regression of genome-wide association studies accounting for ancestry increases power for discovery and improves fine-mapping resolution. Human Molecular Genetics 26, 3639–3650 (2017).

[70] Foley, C. N. et al. A fast and efficient colocalization algorithm for identifying shared genetic risk factors across multiple traits. Nature Communications 12, 764 (2021).

[71] Finucane, H. K. et al. Heritability enrichment of specifically expressed genes identifies disease-relevant tissues and cell types. Nature Genetics 50, 621–629 (2018).

[72] Grotzinger, A. D. et al. Genomic structural equation modelling provides insights into the multivariate genetic architecture of complex traits. Nature Human Behaviour 3, 513–525 (2019).

